# Predicting agranulocytosis in patients treated with clozapine – development and validation of a machine learning algorithm based on 5,550 patients

**DOI:** 10.1101/2025.03.07.25323575

**Authors:** Ebenezer Oloyede, Juan Miguel Lopez Alcaraz, Eromona Whiskey, Olubanke Dzahini, Dan W. Joyce, Sukhi Shergill, Nils Strodthoff, David Taylor, Christian J. Bachmann

## Abstract

**Background:** To prevent clozapine-induced agranulocytosis (CIA), patients’ white blood cell counts are closely monitored, with treatment stopped if the absolute neutrophil count (ANC) drops below 1.5×10^9^/L. While effective, this approach has a high rate of false positives. This study aimed to develop a machine learning (ML) decision-making tool to better predict CIA risk using pattern-based criteria (two consecutive ANCs <0.5×10^9^/L over ≥2 days).

**Methods:** Using a ML technique [gradient-boosted decision trees (GDBT)] we analysed clinical data from 5,550 UK patients treated with clozapine: 2,190 controls with no history of neutropenia and 3,360 cases with at least one neutropenic event, including 358 with pattern-based CIA. Using haematological and demographic data from the current and three prior time windows, predictive models estimated the likelihood of CIA across four time-windows: 1 week, 2 weeks, 1 month, and 3 months respectively in advance. Model performance was evaluated using area under the receiver operator characteristic curve (AUROC), sensitivity, and specificity. We developed another model to predict baseline risk of CIA and compared performance with genetic tests. Explainability analyses identified key features influencing predictions.

**Outcomes:** GDBT models demonstrated strong predictive performance: 1-week forecasting horizon: AUROC 0.99 [95% confidence interval (CI): 0.99–0.99]; 2 weeks: AUROC 0.97 [95% CI: 0.95–0.99]; 1 month: AUROC 0.91 [95% CI: 0.86–0.94]; 3 months: AUROC 0.90 [95% CI: 0.88–0.92]. The baseline model achieved better performance than current genetic tests, with high specificity and sensitivity at varying thresholds. Key discriminative features for CIA included age and baseline haematological values for longer forecasting horizons (1 and 3 months) and current haematological values and treatment duration for shorter horizons (1 and 2 weeks).

**Interpretation:** ML models reliably predict CIA occurrence across short- and long-term horizons, potentially reducing the number of false positives with the current system. Implementation of ML models can reduce unnecessary treatment interruptions and the need for additional blood tests due to suspected agranulocytosis.

**Funding:** The study did not receive direct funding.

**Research in context:** *Evidence before this study:* The only antipsychotic that is effective for treatment-resistant psychosis is clozapine. Tragically, many patients with treatment-resistant psychosis never receive clozapine treatment or receive it many years after “treatment-resistance”. A prominent reason for this is blood tests that are required to detect potential clozapine-induced agranulocytosis (CIA). Despite monitoring being effective, several patients have had to stop clozapine unnecessarily because of the current haematological criteria for discontinuation. In many of these patients, this has resulted in poor clinical and social outcomes. Additionally, many cases of agranulocytosis are identified late under the existing monitoring protocols. At present, there is no reliable way of predicting clozapine-induced agranulocytosis (CIA).

*Added value of this study:* This is the first study to propose that a machine-learning decision tool can reliably predict CIA before it occurs in both the short term and long term.

*Implications of all the available evidence:* Implementation of machine learning algorithms allow prediction of agranulocytosis so that clozapine can be appropriate stopped before it occurs. The algorithm can also prevent unnecessary stopping of clozapine and additional blood testing that is related to spurious blood results.

## 1. Introduction

The reintroduction of clozapine in the 1990s was a paradigm shift for many patients with treatment-resistant schizophrenia (TRS) because of its superior clinical effectiveness compared with traditional antipsychotics. Since the seminal study by Kane et al in the 1980s, numerous studies have demonstrated that clozapine provides improvement in positive symptoms, lower long-term mortality rates, reduced violent behaviour and decreased hospital readmission rates for many patients with TRS [1]. Despite its therapeutic benefits, there is still limited use of clozapine, which has led some clinicians and public health organisations to make efforts to address this issue. One significant barrier to wider utilisation of clozapine is the well-documented risk of agranulocytosis and the consequent need for regular monitoring of neutrophil counts [2].

To prevent CIA-related deaths, current guidelines recommend that if an individual taking clozapine has an absolute neutrophil count (ANC) or white cell count (WCC) below a certain threshold, clozapine must be stopped indefinitely [3]. In some countries, these patients must be registered on a clozapine non-rechallenge database (CNRD), which protects patients from inadvertently being re-exposed to clozapine treatment [4]. Undoubtedly, monitoring schemes have been highly successful in reducing the occurrence of CIA to almost zero. However, the current practice has resulted in many false positives, where patients with mild neutropenia or non-clozapine-related agranulocytosis are erroneously required to discontinue treatment [5]. In many patients, abrupt clozapine discontinuation precipitates a severe relapse that is not alleviated by other antipsychotics, ultimately leading to poor clinical and social outcomes [5].

One of the major challenges in the use of clozapine is the uncertainty surrounding the accurate identification, classification, and prediction of CIA. Current identification is predicated on thresholds of neutrophil counts. Pattern-based criteria are probably more specific to CIA [6]. Using threshold criteria, observational data have suggested that advanced age, female gender, duration of clozapine exposure, and ethnicity are risk factors for CIA. Pattern-based criteria suggest a different array of risk factors [7]. In parallel, genetic risk factors for CIA have low sensitivity, probably because of misclassification bias (i.e. the false association clozapine with severe neutropenic episodes that are unrelated to clozapine). Again, classifying agranulocytosis by the pattern of neutrophil count fall rather than a single below threshold result is likely to reduce the false positive rate of CIA [6]. It is noFigure that genetic variant testing has a high specificity for agranulocytosis, but not high enough to allow the abandoning of blood monitoring in some patients (i.e. those that do not have the variant implicated) [8]. Any system with a specificity approaching 100% would reliably identify those at minimal risk of CIA (so obviating the need for blood monitoring) whereas a system with 100% sensitivity would enable the preclusion of clozapine prescribing in patients who were certain to develop CIA.

To address the difficulties clinicians face in predicting true CIA, our objective was to develop predictive models for CIA that could support clinical decision-making and reduce unnecessary treatment discontinuation. Our hypothesis was that a machine learning (ML) model would outperform current practice and logistic regression in predicting CIA defined by pattern-based criteria.

## 2. Methods

### 2.1. Data source and ethical approval

To accomplish our study aim, non-identifiable data were provided by the two largest manufacturers of clozapine and the associated haematological monitoring services in the UK; Clozaril® (Mylan) monitored by Clozaril Patient Monitoring Service (CPMS) and Zaponex® (Leyden Delta BV) monitored by Zaponex Treatment Access System (ZTAS) [1]. Data included haematological data such as ANC and WCC readings and patient demographic data such as age, ethnicity and gender. The CMPS dataset included national CNRD data on patients who recorded at least one neutropenic reading during clozapine treatment between 2000 to 2021. The ZTAS dataset included patients from one care setting in the UK (South London and Maudsley NHS Foundation Trust) who did or did not experience neutropenia during clozapine treatment between 2004 to 2023. Data on comorbidities or medication usage were not available. Ethical approval was not required according to the UK Health Research Authority (HRA).

### 2.2 Definitions

In the UK, clozapine monitoring is regulated by the Medicines and Healthcare products Regulatory Agency (MHRA), which includes lower discontinuation thresholds for patients with the haematological phenotype benign ethnic neutropenia (BEN) since 2002. A full description of MHRA clozapine regulations is provided elsewhere [3]. Agranulocytosis (severe neutropenia) is typically defined as an absolute neutrophil count (ANC) *<* 0.5 × 10^9^/L. However, under current UK regulations clozapine must be discontinued in the event of an ANC < 1.5 × 10^9^/L and/or a white cell count (WCC) < 3 × 10^9^/L – typically defined as ‘neutropenia’. This recommendation aims to prevent impeding agranulocytosis by detecting early a fall in ANC. Most countries internationally mandate equivalent haematological thresholds for clozapine discontinuation.

Current clozapine monitoring recommendations for CIA result in many false positives, where patients with neutropenia that is mild or not related to clozapine, must discontinue treatment. For example, an isolated ANC < 0.5 × 10^9^/L (i.e., a condition defined as ‘agranulocytosis’) during clozapine treatment may not be related to clozapine at all [9, 10]. To minimise this potential misclassification bias, we defined CIA in our study by ‘pattern-based’ criteria as two consecutive ANC < 0.5 × 10^9^/L over 2 or more days. The ‘neutropenia’ cohort included patients with mild to severe neutropenia (i.e. ANC < 1.5 × 10^9^/L) who did not met criteria for pattern-based agranulocytosis. After combining the CMPS and ZTAS datasets, patients were classified into agranulocytosis (i.e. CIA), neutropenia and control cohorts (i.e. no CIA or neutropenic events).

### 2.3. Study design and participants

A retrospective observational study was conducted combining data from the UK clozapine registries, CPMS and ZTAS. We excluded patients who only had a recorded baseline blood test as this suggested clozapine was not initiated despite registration with the monitoring service. We built a binary classifier, where the positive class denoted a CIA event within a specified forecasting horizon.

### 2.4. Forecasting horizons and datasets preparation

Time frames (i.e. forecast horizon settings) between blood tests were as follows: weekly, two-weekly, monthly, and three-monthly. These horizon settings were selected to represent a variety of monitoring recommendations in national guidelines [4]. In all investigated horizons settings, a series of four measurements were fed into the model, e.g. the monthly horizon setting included today’s measurement and measurements from the three previous months. Each of the four blood measurements were abbreviated and numbered in the order of occurrence i.e. ANC0 at time of prediction, ANC1 one-horizon before, and consecutively up to ANC3. We include a tolerance allowance, where in the absence of this, data was recorded as a missing value - see appendix for detailed information.

Each of the datasets was stratified in 10 folds by patients, ensuring that each fold contained a balanced distribution of unique combinations of gender, season, and diagnosis. The stratification was performed individually for the three cohorts of controls, neutropenia, and agranulocytosis, and corresponding folds were subsequently combined. This yielded 10 folds that matched the respective distribution of the full dataset with respect to gender, season, diagnosis, and neutropenia status as far as possible. Eight folds were adopted for training, one-fold for validation, and one-fold for testing. All hyperparameter details are shown in the appendix.

### 2.5. Statistical analysis, model development, feature selection and explainability

Descriptive analysis of the entire cohort was conducted. Categorical variables were detailed as counts and percentages and continuous variables were described as medians and interquartile ranges (IQRs). The main set of features included ANC, platelet (PLT), and WCC values, the baseline ANC, PLT, and WCC values, demographic data such as gender, the age in years at the prediction time, ethnicity, the season of the year at the prediction time, duration of clozapine treatment and the psychiatric diagnosis (schizophrenia or other). Additional features were engineered from this data. This included percentage changes, rolling means, ratios of the current ANC value against the current and historical PLT, and WCC. Further, we created logarithm, reciprocal and squared values for continuous features such as age, ANC, PLT, and WCC, counts. In total, the model included 104 features, of which 83 were engineered. See appendix Figure 1 for a more detailed description and definition of each feature.

Four main prediction methods for each of the different horizons were investigated.

- First, to simulate current practice (MHRA) standards we predicted CIA from the presence of neutropenia (i.e. ANC0 < 1.5 × 10^9^/L).
- Second, we used a univariate logistic regression model that inputted ANC0 (ANC at prediction time) to simulate MHRA standards from an model perspective.
- Third, we used a gradient-boosted decision tree (GBDT) model (XGBoost). The XGBoost model allows us to better fit the data including missing values, avoiding the drop of samples with missing data or data imputation of missing values.
- Finally, as a model benchmark, we assessed whether the GBDT model provided added value over a multivariate logistic regression model all the features.

As the task contained a heavily imbalanced dataset, each of the training samples were set with a sample weight inversely proportional to class frequencies in the training data. This provided a larger emphasis on the minority classes.

To explore the features that contributed most to a particular forecasting decision, the SHapley Additive exPlanations (SHAP) Treeexplainer [11] was utilised. For a trained model, the explainability algorithm yields a local attribution score, i.e. a relevance value per input feature, which add up to the total model prediction minus the average prediction. In our study, “positive correlation” indicated an increased probability of CIA as the predictor value decreases and “negative correlation” indicate a reduced probability of CIA as the predictor value increases. A summary of the model behaviour across all samples in terms of mean (absolute) SHAP values is provided in the appendix. All computations were performed using Python V.3.10 (Python software foundation, http://www.python.org) and the scikit-learn package V.1.1.3. Figure 1 provides a summary of the model development.

**Figure 1.**
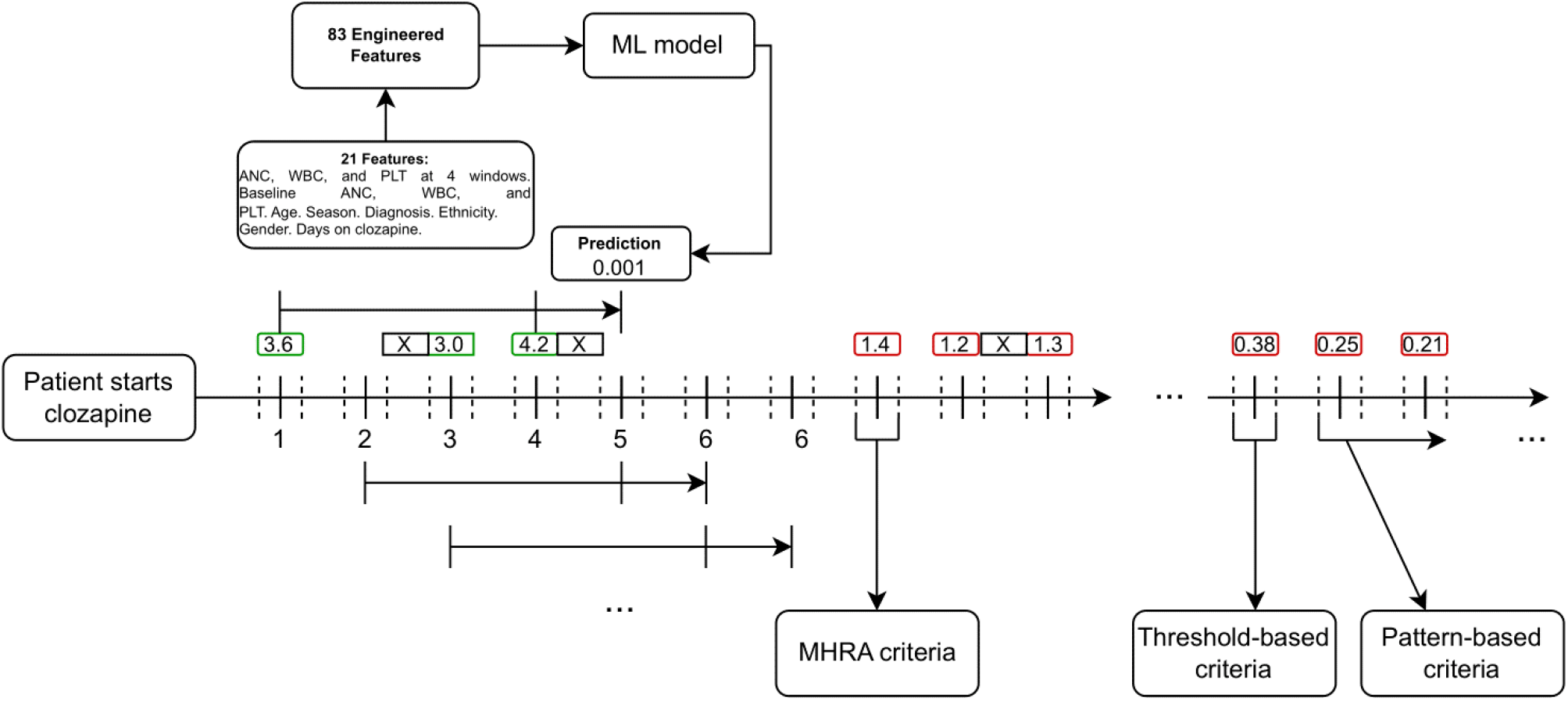
Visual representation of the proposed sampling and modelling pipeline. ANC values are provided in green and red boxes for illustrative purposes, where green represent the data used for training up to ≥ 1.5 × 10^9^/L and red represents the possibility of CIA occurrence. We considered fixed time horizons e.g. week 1, 2, and so on, denoted as solid lines above each number in the horizon. We also included tolerance thresholds for haematological measurements usage denoted as dashed lines before and after solid lines, where absent data was not included as a predictor, denoted by ‘X’. Our machine learning model operates using 4 forecasting blocks to predict one block ahead. The 4 blocks contain 21 features of haematological and demographic data and a further 83 engineered features. The MHRA criteria represents the < 1.5 × 10^9^/L threshold that is used in clinical practice to predict CIA. Threshold-based criteria represents the clinical definition of agranulocytosis and pattern-based criteria represents the CIA definition we adopted.

### 2.6 Performance metrics

Models to predict CIA were evaluated with the area under the receiver operating characteristic curve (AUROC); sensitivity and specificity in the neutropenia cohort, and specificity in the control cohort. Predictive binary tasks rely on confusion matrix elements such as true positive (TP), true negative (TN), false positive (FP), and false negative (FN). To convert the continuous probabilities to binary predictions, we selected a threshold that balances TPs and TNs at a desired design choice. We applied a model selection strategy based on AUROC performance on the validation set (fold 9). Test sensitivities were controlled at different threshold values (e.g., 0.90, 0.85, 0.80). When the threshold achieved the desired sensitivity, we compared specificities. AUROC, sensitivity, and specificity for the neutropenia and agranulocytosis cohorts were reported using the test sets confusion matrix (fold 10). For control patients, specificity was reported as TPs are not present. Empirical bootstrapping (n=1000 iterations) was used to obtain 95% confidence intervals (CI). We applied the sensitivities thresholds on the lower CI bound from the test set to ensure conservative performance. The calibration of the XGBoost models were evaluated using calibration curves and compared with the multivariate logistic regression model using the expected calibration error (see appendix).

### 2.7. Baseline prediction model

As a secondary objective, we investigated the feasibility of predicting a patients’ baseline risk of CIA (i.e. before clozapine initiation) using an XGBoost classifier. The model included previously described hyperparameters, patient demographic information, baseline blood counts (ANC, WCC and PLT) and the engineered features. In total, 30 features were included. We aimed to compare this model performance with existing baseline prediction models in the form of genetic predictors for CIA, in terms of sensitivity and specificity.

## 3. Results

### 3.1 Baseline characteristics

The dataset consisted of 414,312 unique blood measurements from 5,550 patients after excluding 78 patients who only had a baseline blood count. Figure 2 provides a comprehensive description of the study cohort. A total of 3,002 patients had a neutropenic event - the median age of this group was 38 years (IQR 20), 62% were male and the median duration of clozapine treatment was 2 (IQR 5). Over a median follow-up of 4.73 (5.51) years, 358 patients developed pattern-defined agranulocytosis. The median age of this cohort was 47 years (IQR 18), 63% were male and the median duration of clozapine treatment was 1 year (IQR 2). There were 2,190 controls, patients who had no neutropenia or agranulocytosis while receiving clozapine treatment - the median age of this group was 37 years (IQR 18), 63% were male and the median duration of clozapine treatment was 6 years (IQR 13).

**Figure 2.**
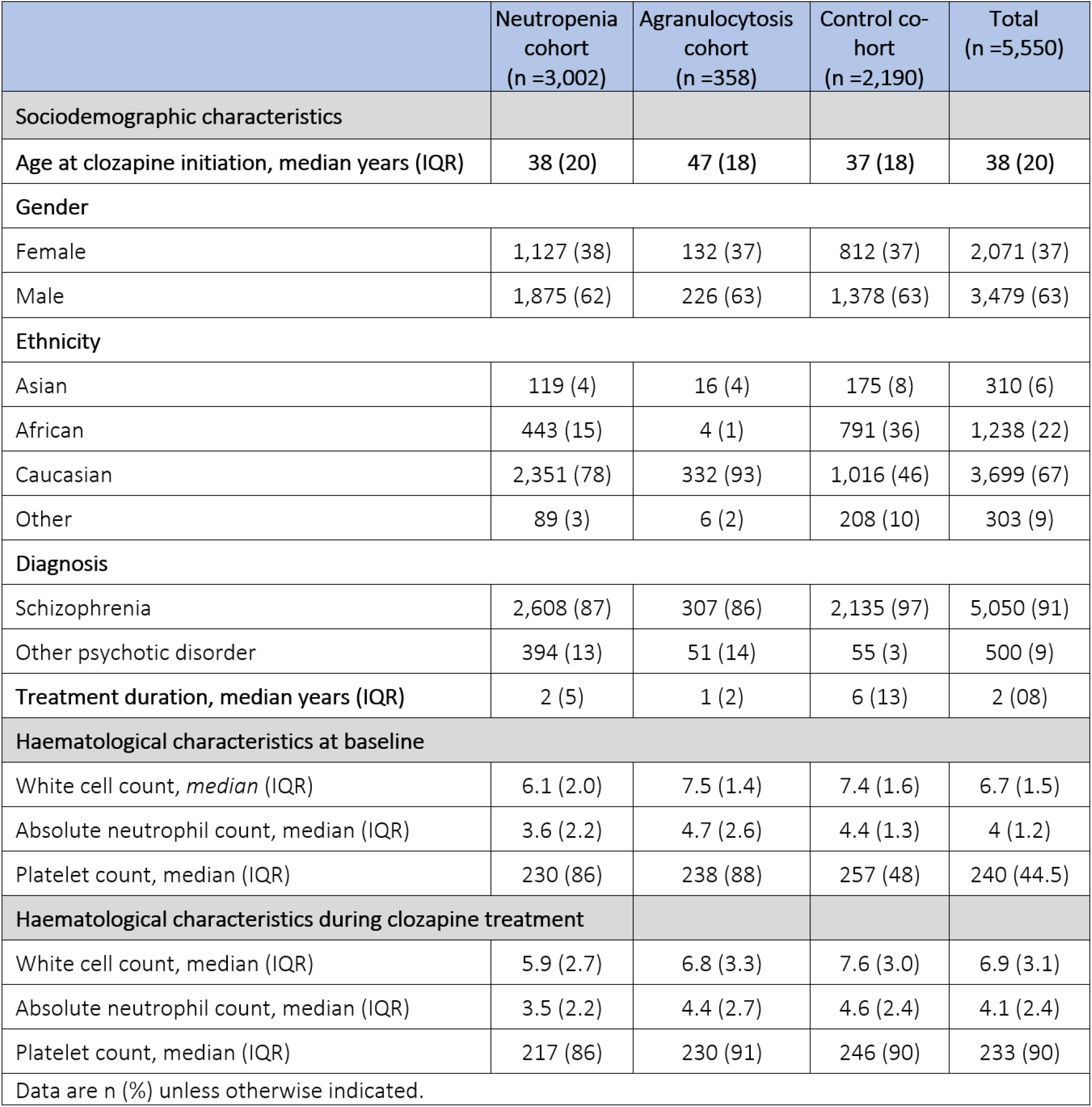
Performance metrics for the various models.

### 3.2 Predictive performance

Figure 3 shows the AUROC values and the associated 95% CI for the univariate logistic regression (using a single ANC count) and XGBoost models across the different forecasting horizons. In addition, the sensitivities and specificities of current MHRA standards for predicting CIA are presented in Figure 4. As a comparison, the specificities of current monitoring standards exceed 0.90 for all forecasting horizons and sensitivities range from 0.08-0.10. For the univariate logistic regression model (single ANC count), the AUROCs were 0.69 for 1 month and 0.52 for 3-month horizons. For shorter horizons of 1 week and 2 weeks, the univariate logistic regression (single ANC count) AUROC values were 0.96 and 0.83 respectively. For all the settings, the XGBoost model had high performance, particuarly for the longer-term, 1 month and 3-month, horizon forecasts. The AUROC for XGBoost models exceeded 0.90 for all forecasting horizons. The calibration plots, calibration error and AUROC demonstrate better performance for the XGBoost models across different forecasting horizons, outperforming univariate and multivariate (with all features) logistic regression models (see appendix).

**Figure 3.**
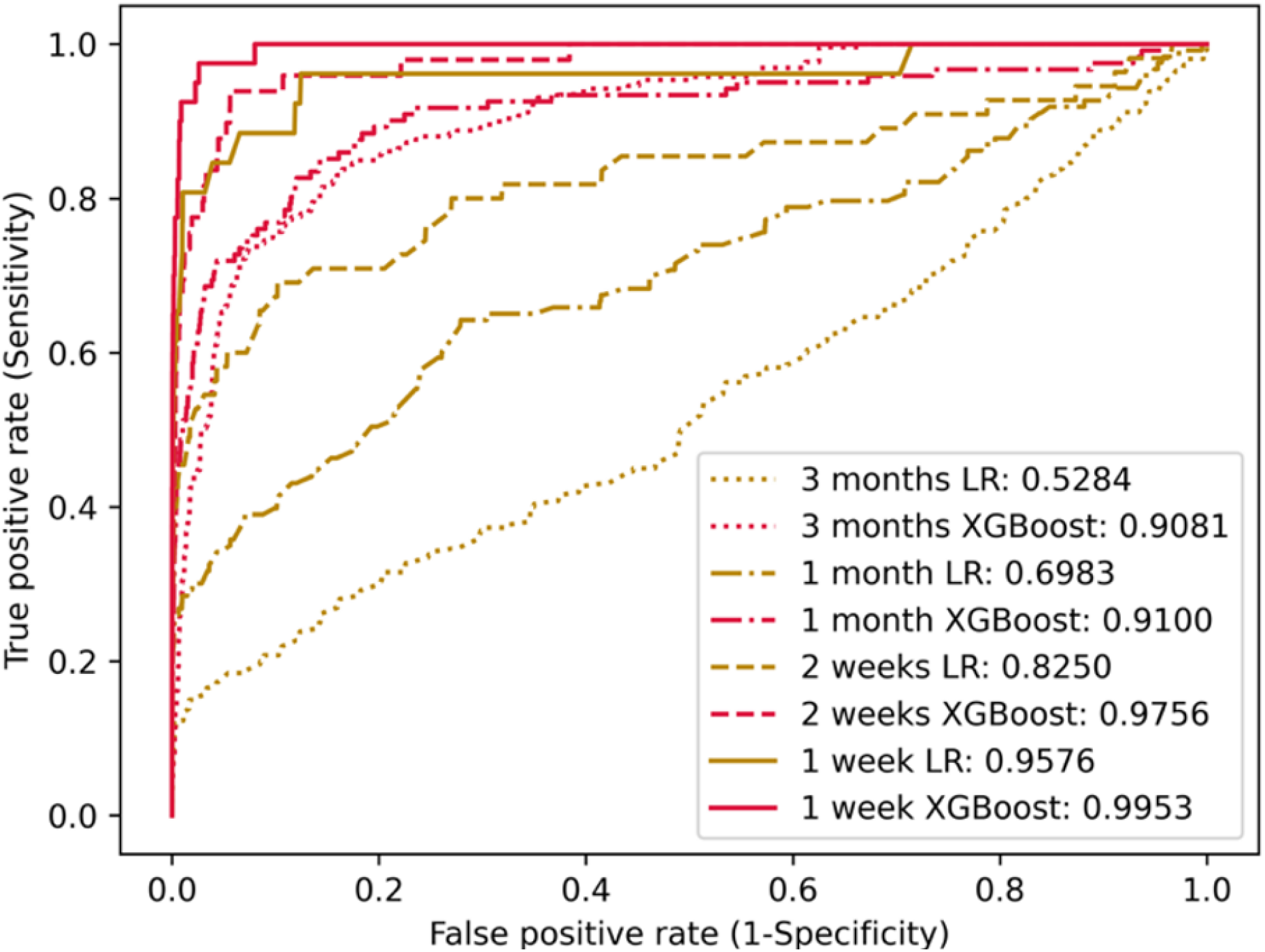
AUROC curves for the ML and univariate logistic regression models (single ANC count) at different forecasting horizons.

**Figure 4.**
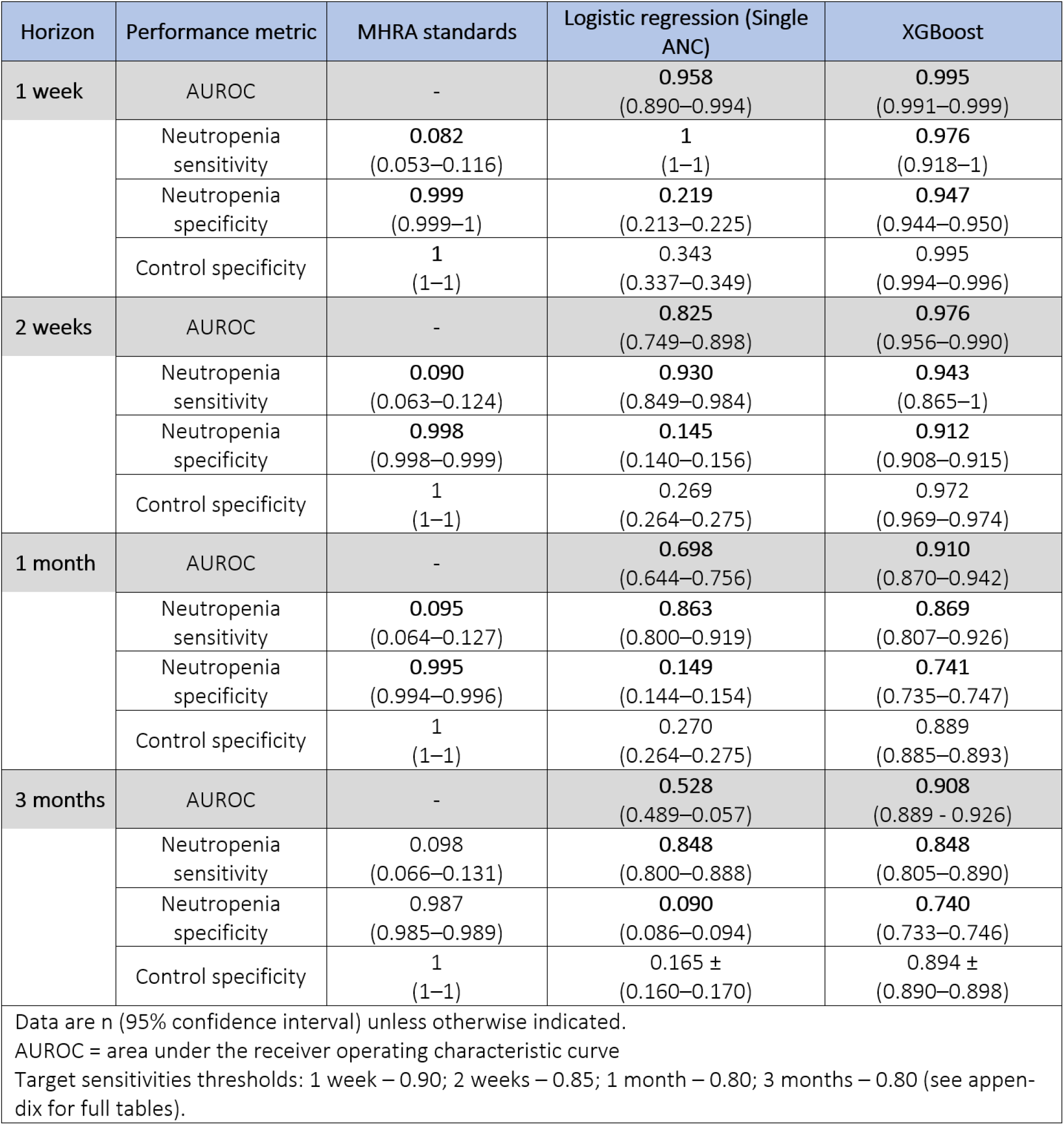
Performance metrics for the various models.

### 3.3. Baseline prediction model

Figure 5 contains the predictive performance for the baseline prediction model. Overall, we achieve an AUROC of 0.655 (0.642-0.667). The baseline model achieves comparable performance to existing genetic predictors. Figure 23 in the appendix provides specific performance at different specificities. Figure 24 in appendix contains more sensitivity thresholds.

**Figure 5.**
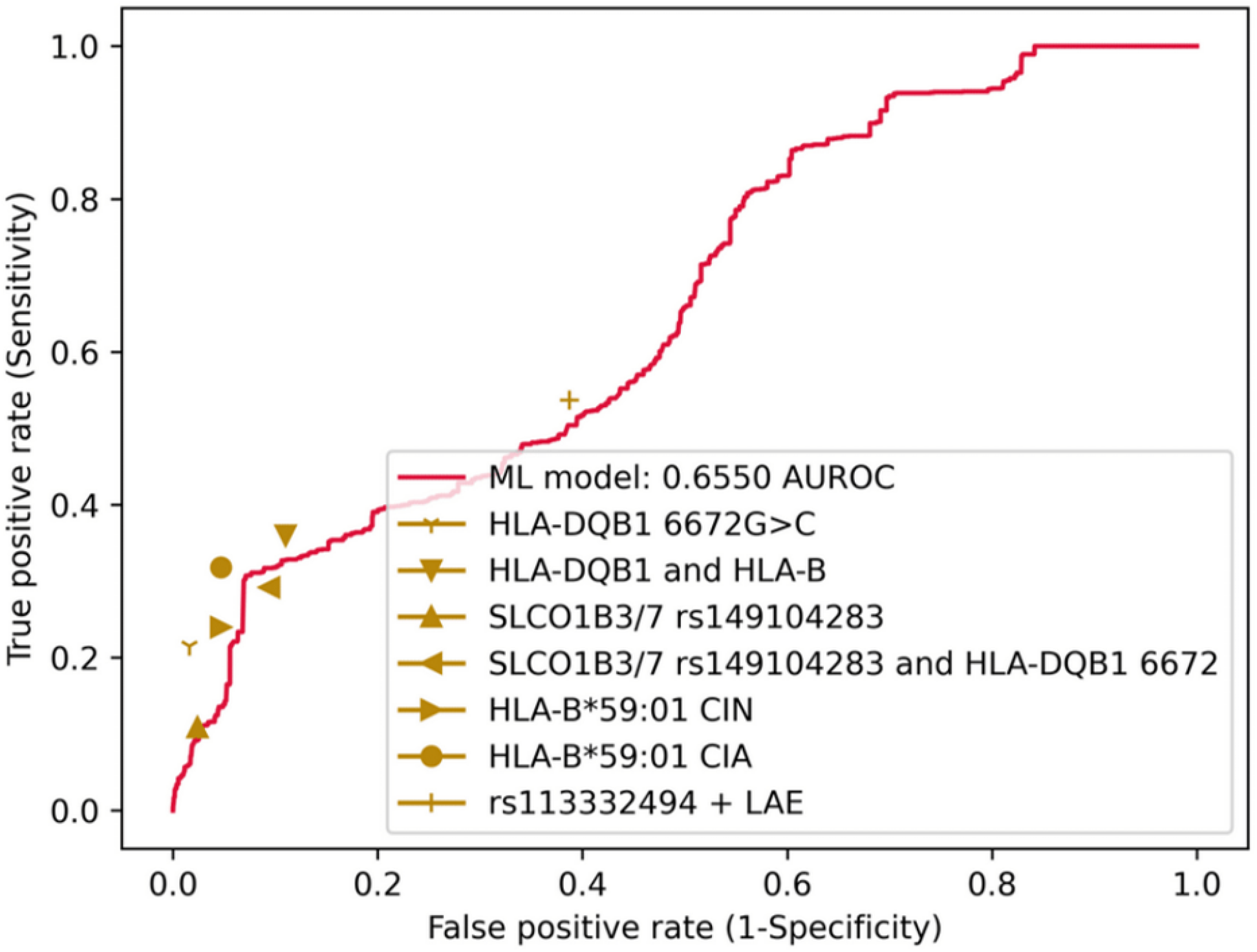
AUROC curves (in red) for the baseline ML prediction tool and the sensitivity and specificity values achieved by genetic predictors (in gold), where routine threshold-based definition of agranulocytosis was adopted in the ML model.

### 3.4. Model explainability

Figure 6 presents the top seven model variables in decreasing magnitude of contribution at the different forecasting horizons and baseline prediction model. The last column shows the number and contribution of the other variables. Generally, across all forecasting periods, the duration of clozapine use demonstrated a negative correlation with CIA risk (i.e. reduced risk of CIA as duration of clozapine use increases). Caucasian race demonstrated positive correlation with CIA risk, whereas Black race demonstrate negative correlation. Haematological readings at prediction time were more significant at shorter horizons (1 week, 2 weeks). Discriminative features for the baseline prediction tool were like the forecasting models. Additional features included baseline ANC count, and the age to haematological reading ratio at baseline. More detail on model explainability including SHAP plots for each of the forecasting horizons and the ML baseline prediction can be found in the appendix.

**Figure 6.**
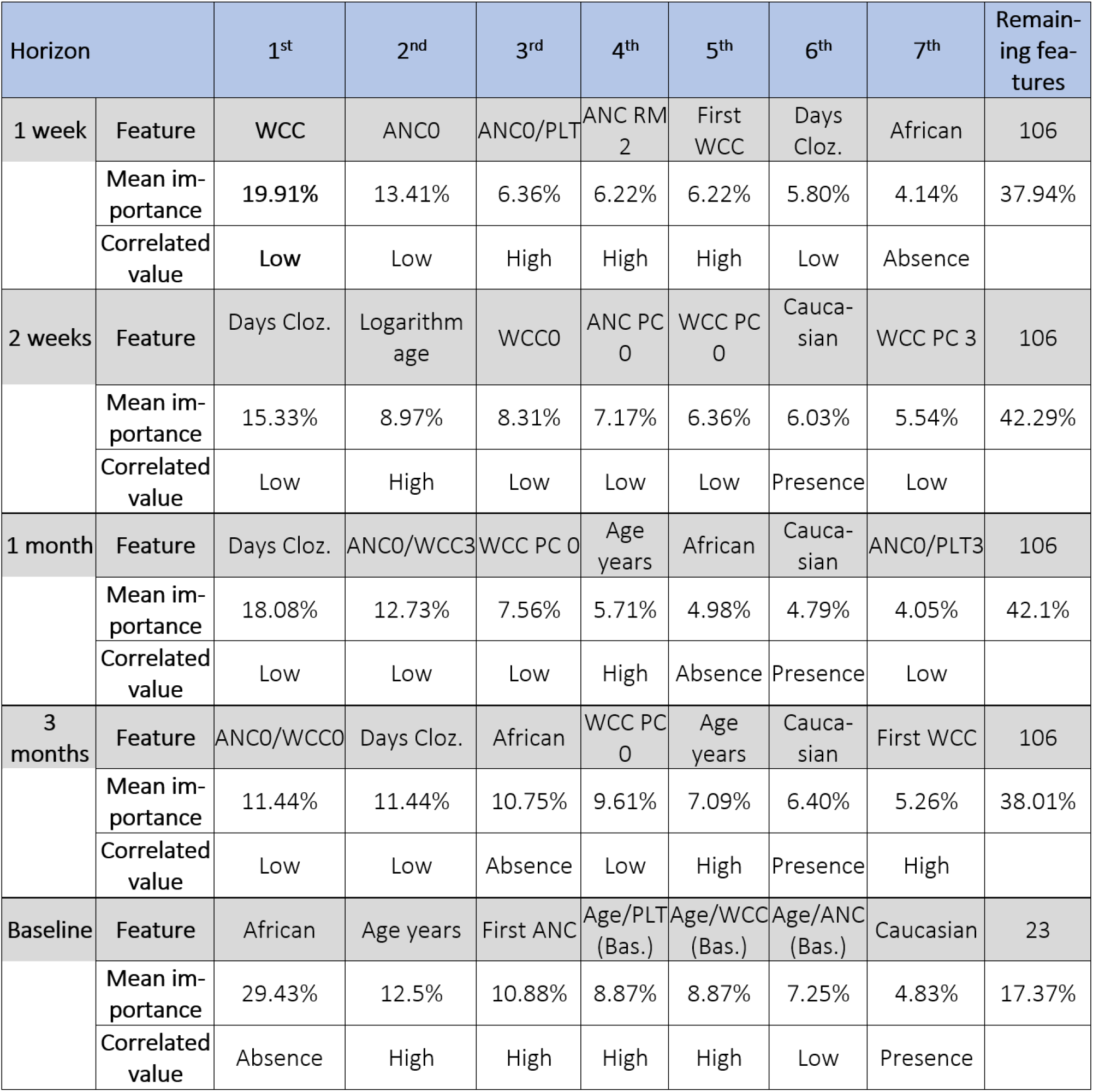
The top seven predictors for each of the investigated horizons, and for the baseline prediction tool. The last column indicates the sum importance of the remaining predictors. Individual rows present the predictors name, mean importance, and specific values that correlate with CIA risk. For continuous variables, both low and high ranges are considered, while binary values indicate the presence or absence of the predictor.

**Figure 7.**
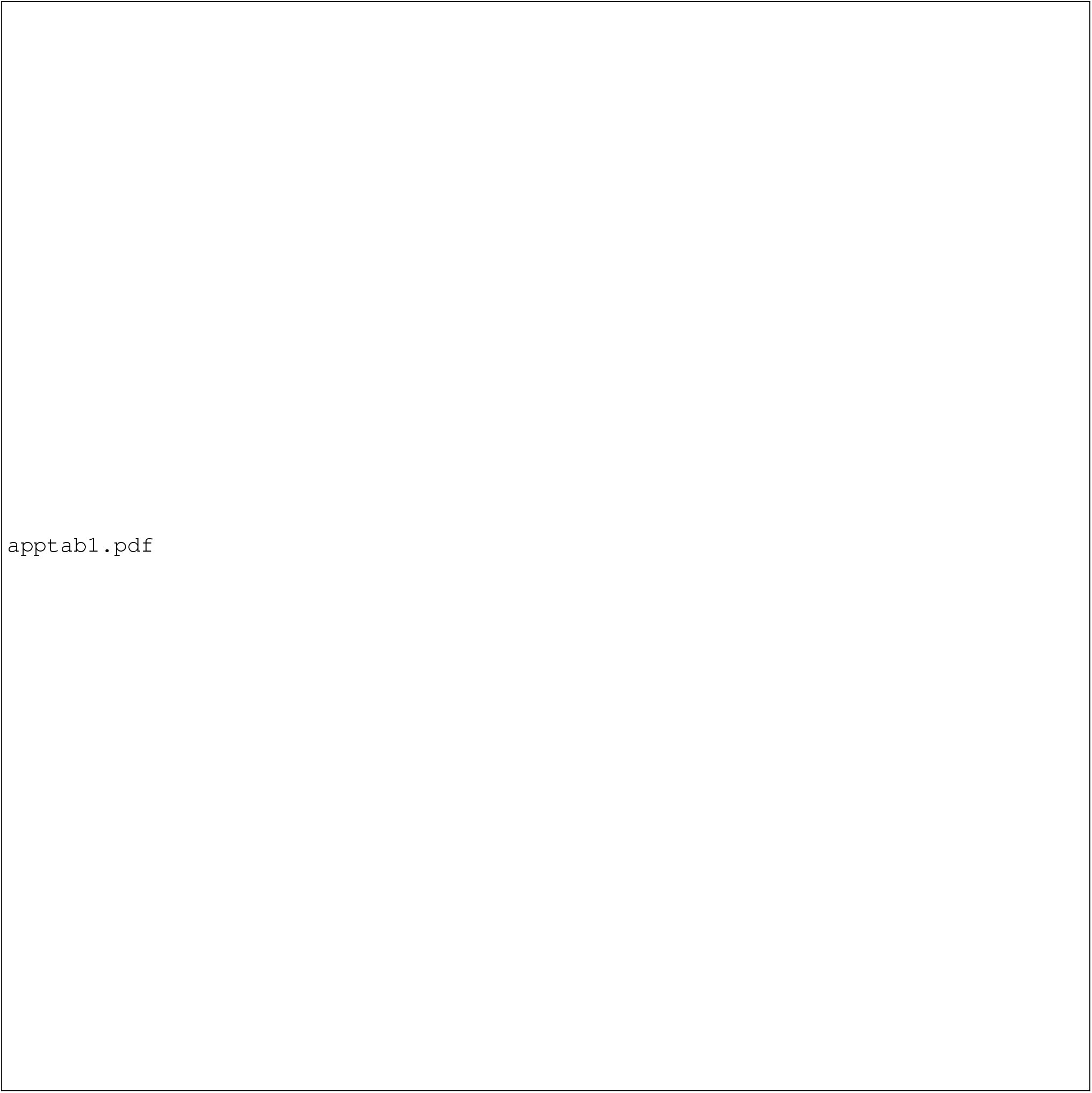
Hypothetical observation period.

## 4. Discussion

### 4.1. Summary of findings

In this cohort study, we developed the first ML model (XGBoost) that can be used alongside standard monitoring to better predict CIA occurrence with high sensitivity and specificity at various time horizons. Its sensitivity is comparable to standard monitoring and higher than genetic predictors. Its specificity is considerably higher than standard monitoring and similar to baseline genetic predictors. As such, our model represents a major step forward in assuring the safe use of clozapine and in precluding the unnecessary discontinuation of clozapine.

### 4.2. Model performance

As a risk mitigation strategy for CIA, most clinical guidelines dictate that patients must discontinue clozapine if the ANC <1.5 × 10^9^/L. However, the rationale for this discontinuation criterion continues to be questioned. Several lines of evidence suggest that the current criteria for clozapine discontinuation impose unnecessary restrictions on clozapine use and can be safely relaxed without compromising patient safety [3, 12]. Our study revealed that current MHRA discontinuation criteria have a sensitivity of 0.0824 (0.0528 - 0.1156) and a specificity of 0.9993 (0.9989 - 0.9997) at the one-week horizon. In other words, when predicting pattern-based agranulocy-tosis within one-week of the first <1.5 × 10^9^/L, currently up to 90% of blood samples are not predicted to have CIA when they do. This observation is somewhat surprising given the prevailing belief that current monitoring effectively identifies most cases of agranulocytosis. However, our findings can be better understood in light of our recent research, which indicated that 70% of patients who develop agranulocytosis do so from a normal absolute neutrophil count (ANC), without any prior documentation of mild to moderate neutropenia [7]. Moreover, it is plausible that some cases of agranulocytosis occur after one week of the initial ANC <1.5 × 10^9^/L, explaining the marginally better sensitivities achieved in the longer time window models.

Given the negative clinical and economic consequences associated with the discontinuation of clozapine, a critical question arises: is there a more efficient method for identifying clozapine-induced agranulocytosis (CIA) ML is increasingly recognised as a valuable tool for supporting clinical decisions across various medical fields. Our analysis consistently demonstrated that the ML model based on the XGBoost algorithm provided superior predictive performance for CIA compared to both current practices and logistic regression. Specifically, at a one-week forecasting horizon, the XGBoost model achieved an AU-ROC of 0.9953 (95% CI 0.9907–0.9985), with a sensitivity of 0.9762 (95% CI: 0.9183–1.0000) and a specificity of 0.9474 (95% CI: 0.9442–0.9504). Furthermore, our ML model outperformed the multivariate logistic regression model, as indicated by the AUROC values (see Figure 8 in the appendix).

**Figure 8.**
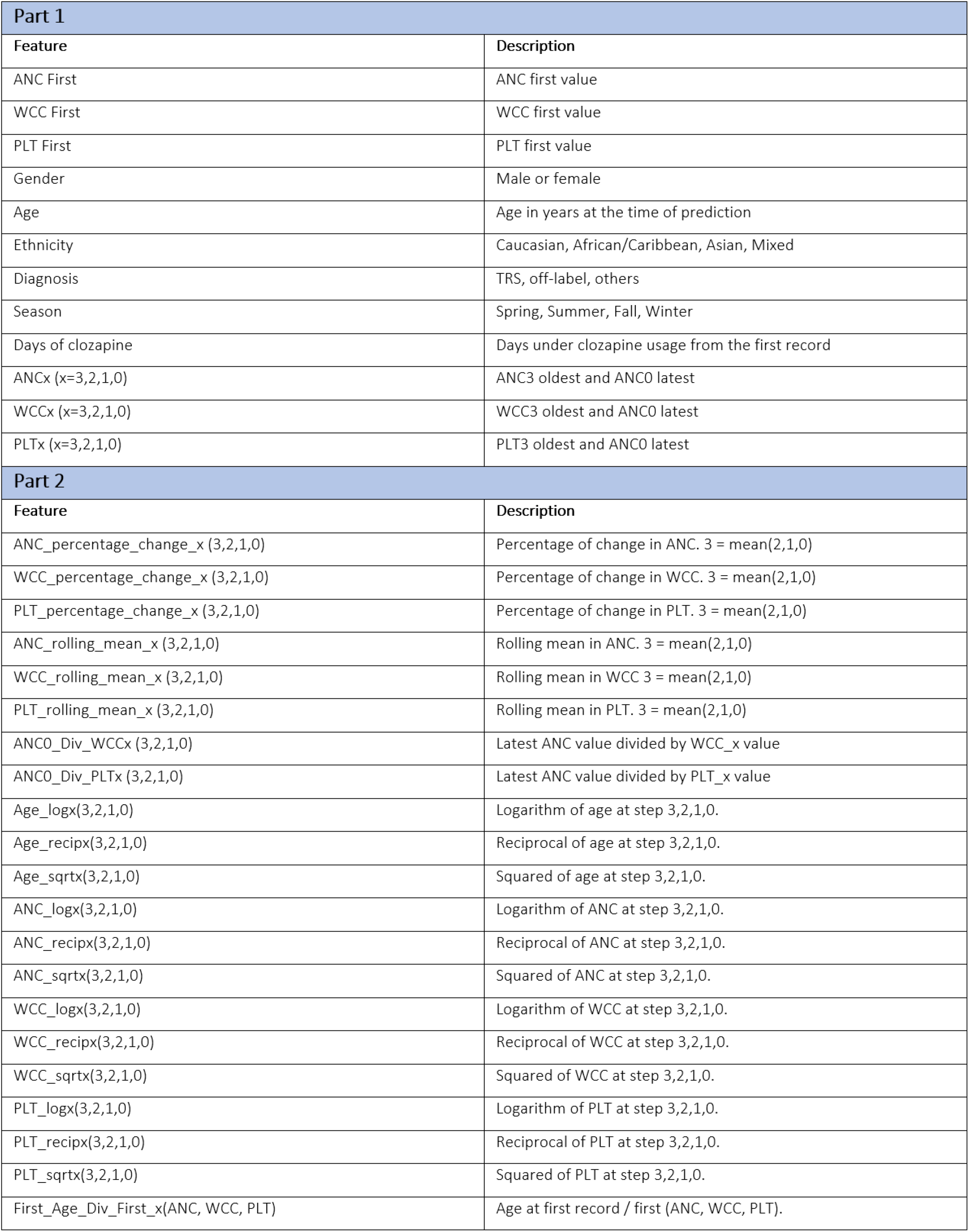
Model features set.

**Figure 9.**
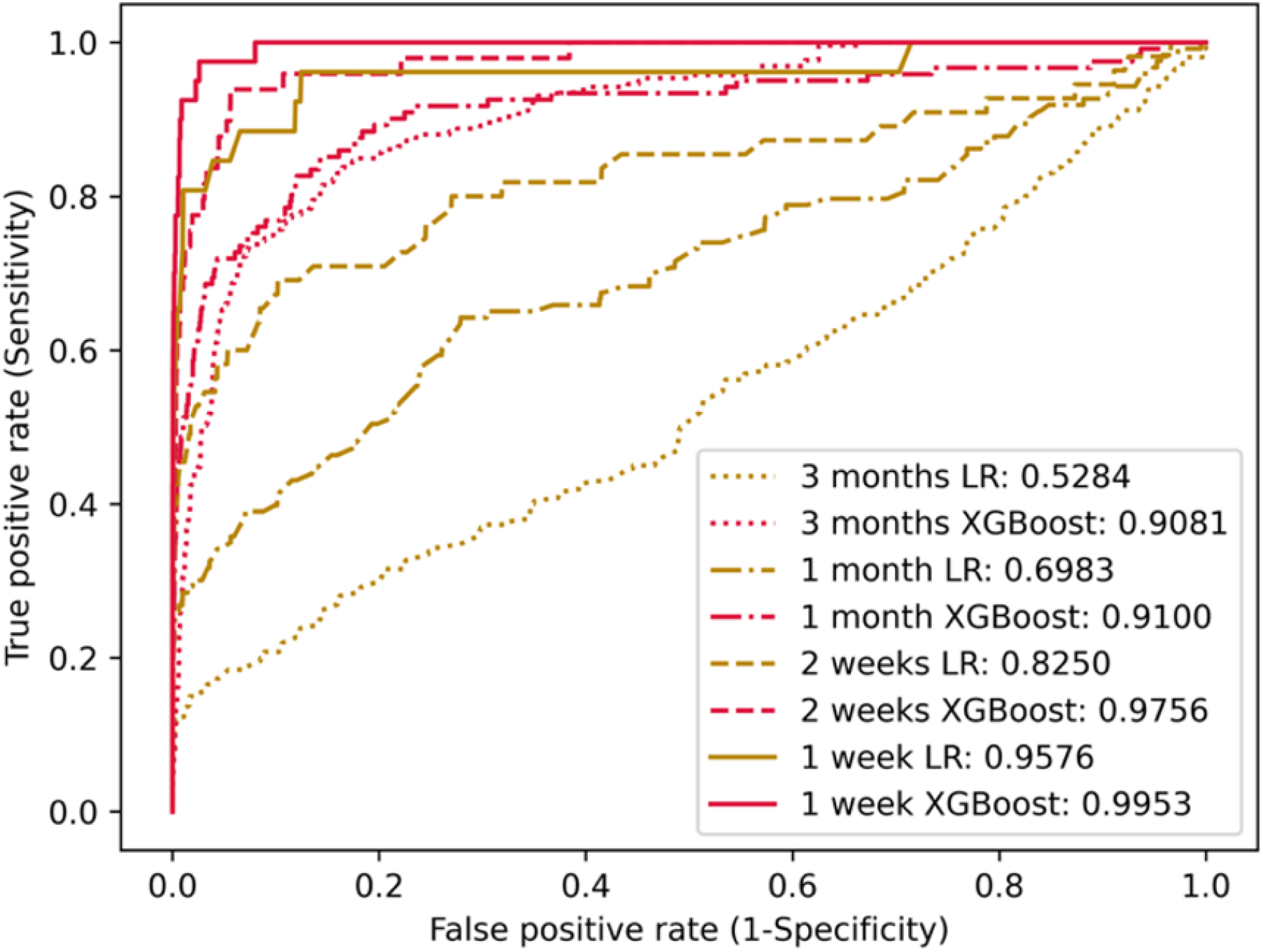
AUROC curves for the ML and univariate logistic regression models (single ANC count) at different forecasting horizons

So, how could this affect practice? Assuming 10,000 patients prescribed clozapine are treated and monitored according to current protocols, approximately 380 individuals would have neutropenia and 40 will develop threshold-defined CIA [13]. Based on a the most recent blood test, MHRA standards correctly classify all those without CIA. However, the ML tool incorrectly classifies 18 individuals as having CIA when they do not. In the patients who recorded neutropenia, MHRA standards accurately identify 340 individuals as not having CIA while the ML tool identifies 322. The most noFigure difference between the ML and MHRA standards is in the identification of true CIA case: the ML tool identifies only 3 patients with CIA, while the MHRA standards correctly identify 39 cases (See Figure 27 in the appendix).

### 4.3 Feature importance

The past decade has seen numerous attempts to identify predictors for CIA. However, existing studies have been limited by small datasets and simplistic analytic approaches, resulting in conflicting conclusions. Thus far, risk factors suggested in the literature include increasing age, female gender, specific human leucocyte antigen (HLA) haplotypes and the duration of exposure to clozapine [14–21]. Among the key features in our prediction model, some have been previously identified, while others are newly discovered influencing variables. For instance, a recent retrospective cohort study of 520 patients by Johannsen et al. found that low baseline ANC (OR 24.4; 95% CI 6.4–215.0, p < 0.001) and a history of previous neutropenia episodes (OR 11.4; 95% CI 3.9–34.9, p < 0.001) were risk factors for mild to severe neutropenia [15]. In contrast, a recent study of 1,038 patients prescribed clozapine in Brazil, observed that only the presence of a severe medical comorbidity such as HIV was a risk factor for moderate to severe neutropenia (HR 69.4; 95% CI 37.5–128.4, p < 0.001) [22].

Although direct comparisons are challenging due to differences in the features and categories of neutropenia studied, our analysis identified duration of clozapine treatment, ethnicity and age as prominent features across all four forecasting horizons. Unlike previous research, our study did not observe sex to be a predictor for CIA, suggesting that it may not significantly contribute to an individual’s risk or that the risk can be explained through other observed clinical characteristics. Interestingly, our finding of a reduced risk of agranulocytosis in patients of black ethnicity is not a newfound phenomenon. As early as 1958, observational data from Pisciotta et al. suggested a lower susceptibility to agranulocytosis in black patients treated with phenothiazine derivatives such as chlorpromazine [23]. However, misconceptions among clinicians have persisted, leading to reduced use of clozapine in this population [24]. It can be argued that one of the most influential factors contributing to this is clinicians misinterpreting non-pathologically lower ANC (i.e. BEN) as CIA [9].

Notably, one of the aforementioned studies proposed that white ethnicity (HR 0.53; 95% CI 0.29–0.99, p < 0.05) could be a protective factor for clozapine-associated moderate to severe neutropenia [22]. However, this finding has at least three major flaws. First, the authors did not differentiate between moderate and severe neutropenia, despite the evidence suggesting that clozapine primarily causes severe neutropenia (i.e., agranulocytosis) [6]. Second, the study results are likely biased as black patients are more likely to experience moderate neutropenia due to BEN. Third, there has historically been considerably more admixture in the country sampled, compromising the reliability of self-reported ethnicity as a measure.

Several new data-driven features were identified in our study, such as the percentage change of WCC or ANC, which align with a proposed immunological basis for drug-induced neutropenia that is characterised by a rapid decline in neutrophil counts [25–27]. Other newly discovered features, such as ANC divided by WCC at different time points, are not easily explained but may signal a general reduction in the ratio of neutrophil to total WCC over time in individuals who develop CIA.

### 4.4. Baseline prediction model

The goal of clozapine initiation in individuals with TRS is to achieve a good clinical response without any serious adverse drug reactions (ADR), including CIA. To achieve this, preventative strategies based on robust baseline risk assessments are crucial. Considering this, we aimed to develop an ML model to identify the baseline risk of CIA using routine demographic and clinical data. To date, efforts to identify baseline risk for CIA have primarily focused on genetic data. However, genetic prediction for the CIA remains more of an aspiration than a reality. A recent review described optimal sensitivities and specificities of different genetic variants as 0.215 and 0.984 respectively (HLA-DQB1 6672G¿C) [28]. Whereas a more recent study in individuals of European ancestry achieved a sensitivity and specificity of 0.5370 and 0.6129, respectively [8].

While beyond the scope of the present study, the underachievement of genetic models may be due to misclassification bias, where patients with recorded ANC <0.5 × 10^9^/L unrelated to clozapine are recorded and analysed as such. As indexed by sensitivity and specificity values, our model performed substantially better than genetic predictors, while highlighting ethnicity, age, baseline ANC, and various ANC and PLT measurements as the most discriminative features. This was notably even with the routine threshold-based definition for agranulocytosis. This could be because each of the genes is linked to a specific demographic, limiting its applicability in the general population. Future inclusion of genetic data to our baseline prediction tool could plausibly improve model performance. From a practical perspective, our ML model is likely more cost effective potentially widening its applicability in clinical practice.

### 4.5. Clinical implications

In many patients with TRS, continuation of clozapine is the only route to achieving long-term clinical stability. Although current monitoring schemes help to prevent CIA and its complications, they also result in several false positives, leading to psychiatric decompensation and poor prognosis for many patients [3, 7]. Identifying true cases of CIA is currently a complex and resource-intensive task. Therefore, an automated early warning system that can predict and rule out CIA alongside blood monitoring would offer numerous benefits. From an implementation standpoint, since demographic and haematological data are readily available in EHR systems and easily extracFigure, a model based on this data could be more easily adopted by healthcare providers on an international scale.

Integrating our model into healthcare systems and clozapine monitoring databases could provide real-time risk scores to patients, for instance, during point-of-care ANC monitoring. Furthermore, a CIA prediction tool could improve patient satisfaction and save significant healthcare resources by reducing monitoring requirements for patients at low risk for CIA. From a safety standpoint, patients who are deemed as higher risk of developing CIA could receive enhanced monitoring or appropriate discontinuation of treatment, moving away from a “one-size-fits-all” approach to tailored screening strategies. Another benefit of such a tool would be a potential reduction in polypharmacy and pill burden from unnecessary CIA mitigation strategies such as granulocytecolony stimulating factor (G-CSF) and lithium, further improving patient acceptance and reducing non-compliance. Given the ongoing challenge of changing clinicians’ perspectives on clozapine’s risk, efforts may be better focused on implementing enhanced decision support tools such as ours to alleviate clinicians concerns and improve confidence in prescribing clozapine. In further research, we plan to validate our model using external data.

### 4.6. Strengths, limitations and future work

Our study has several strengths. Our cohort is large and includes a highly diverse population, increasing the generalisability and clinical application of the model. Additionally, a key strength of our study lies in our definition of CIA, which aligns with accumulating data describing CIA as a distinct pattern of continuous and rapid decline in neutrophil count to zero or near zero [6]. Another strength of our study is the innovative use of ML techniques to utilise the abundant data in the electronic medical record. Unlike traditional risk prediction models that use data from a single time point and therefore only incorporate a small portion of the available patient data, our algorithm utilises comprehensive time-series data.

Nevertheless, there are several limitations that merit consideration. First, while it is planned future work, our model has not been externally validated using national or international databases, which limits statements about the generalisability. Second, although our study included a variety of features, there were other potential features, such as concomitant medication and physical health comorbidities that were not included in our model. Yet, considering the performance of our model and how readily available the included features are, it is plausible that our model can optimise the identification of CIA on a wide scale. Lastly, we acknowledge that, like any retrospective study using observational data, we cannot establish causation of the observed risk factors or rule out the significance of unobserved factors. However, the proposed final model provides highly accurate characterisations of patient risk based on the included features.

### 4.7. Authors contribution statement

J.M.L.A. and N.S. conceptualized and designed the methodology and experiments. J.M.L.A. conducted the experiments, while both contributed to the planning and refinement of the methods. E.O., E.W., O.D., D.W.J., S.S., D.T., and C.J.B. provided clinical expertise and contributed to the interpretation of results. E.O. facilitated access to the dataset. J.M.L.A. and E.O. wrote the first draft, which was revised by N.S., D.T., and C.J.B. All authors reviewed and approved the final manuscript.

### 4.8. Data sharing statement

The data that support the findings of this study are available from Viatris. Restrictions apply to the availability of these data, which were used under license for this study.

## Data Availability

The data used in this study are not publicly available due to patient confidentiality but can be accessed upon reasonable request to the authors and subject to relevant ethical approvals.

## Appendix

### Hypothetical observation periods and model features

Feature sets were divided into two – part 1 consists of original features, and part 2 consists of derived features. These features encompass information such as gender, age, ethnicity, diagnosis, season, days of clozapine usage, various blood cell counts, percentage changes, rolling means, logarithmic, reciprocal, and squared values, and ratios of different parameters related to medical records and health conditions.

#### Exploratory data analysis

Exploratory data analysis revealed the following insights:

- Asians with CIA events report a lower median baseline ANC, WCC, and PLT value (3.9, 6.975, 238.5) compared to Asians without CIA events (4.2, 7.1, 251). Other ethnic groups demonstrate an inverse relationship.
- Males with CIA events report a lower age per ANC at first measurement time (8.94) compared to males without CIA event (8.96). This behaviour occurs only in males and is specific to ANC.

#### Model performances

Figure 10 presents the overall predictive performance for the three investigated models; logistic regression with a single ANC value which aims to mimic MHRA standards, logistic regression with all the investigated features which aims to improve MHRA standards from a linear perspective, and XGBoost with all the investigated features which aims to improve MHRA standards from a non-linear perspective. Firstly, comparing the two logistic regression models we observe that the model with all features outperformed the one with single feature in the long horizon settings (3 months and 1 month). Notably, the single feature model outperformed the model with all features on shorter horizon settings (2 weeks and 1 week). This may be related to samples being dropped from the logistic regression model with all features as logistic regression does not handle missing data. Secondly, when comparing logistic regression and XGBoost with all features, we observed that XGBoost outperformed logistic regression in all investigated horizons. For the XGBoost after performing an hyperparameter search, we found that trees with a maximum depth of 1 yielded better outcome. The datasets were standardised only for the logistic regression models.

**Figure 10.**
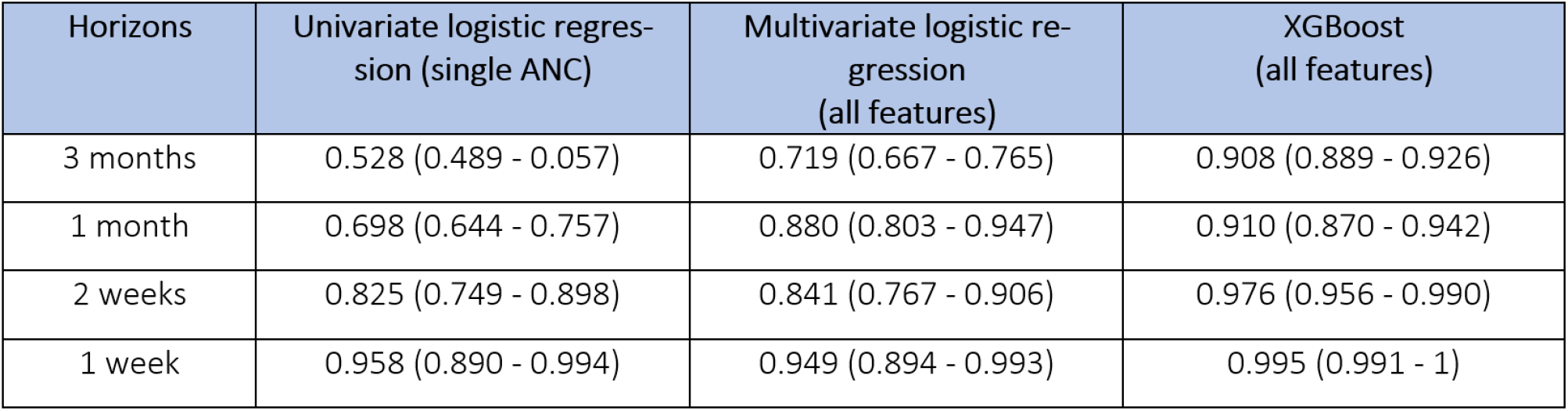
Predictive performance comparison between logistic regression with a single ANC count, logistic regression with all the investigated features, and XGBoost with all the investigated features

#### 1 week forecasting horizon

Figure 11 displays the results for the weekly settings over the investigated sensitivity thresholds for the three investigated predictive models.

**Figure 11.**
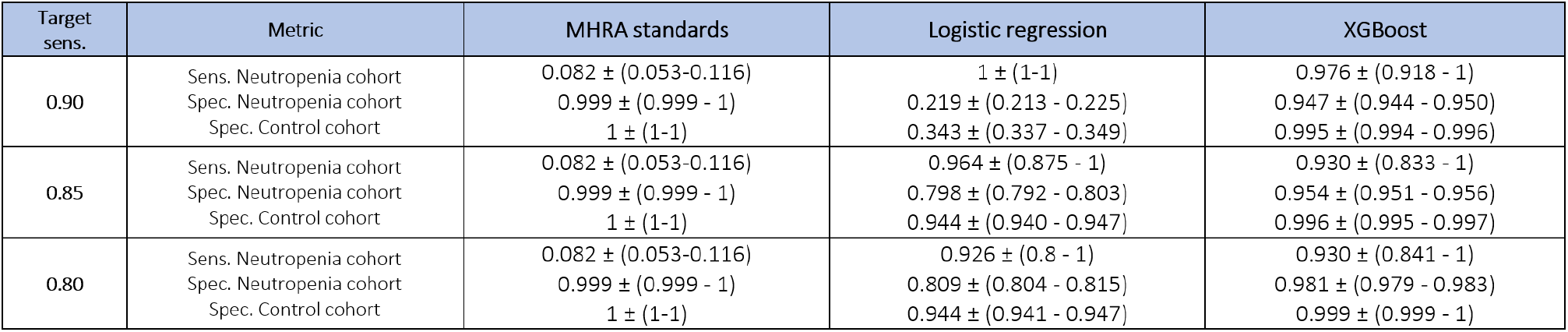
Predictive performance: 1 week forecasting horizon sensitivities and specificities

Figure 12 contains the SHAP values for the weekly model. Overall, and by order of importance, we can observe some of the features that are more likely to be associated with an event of CIA:

**Figure 12.**
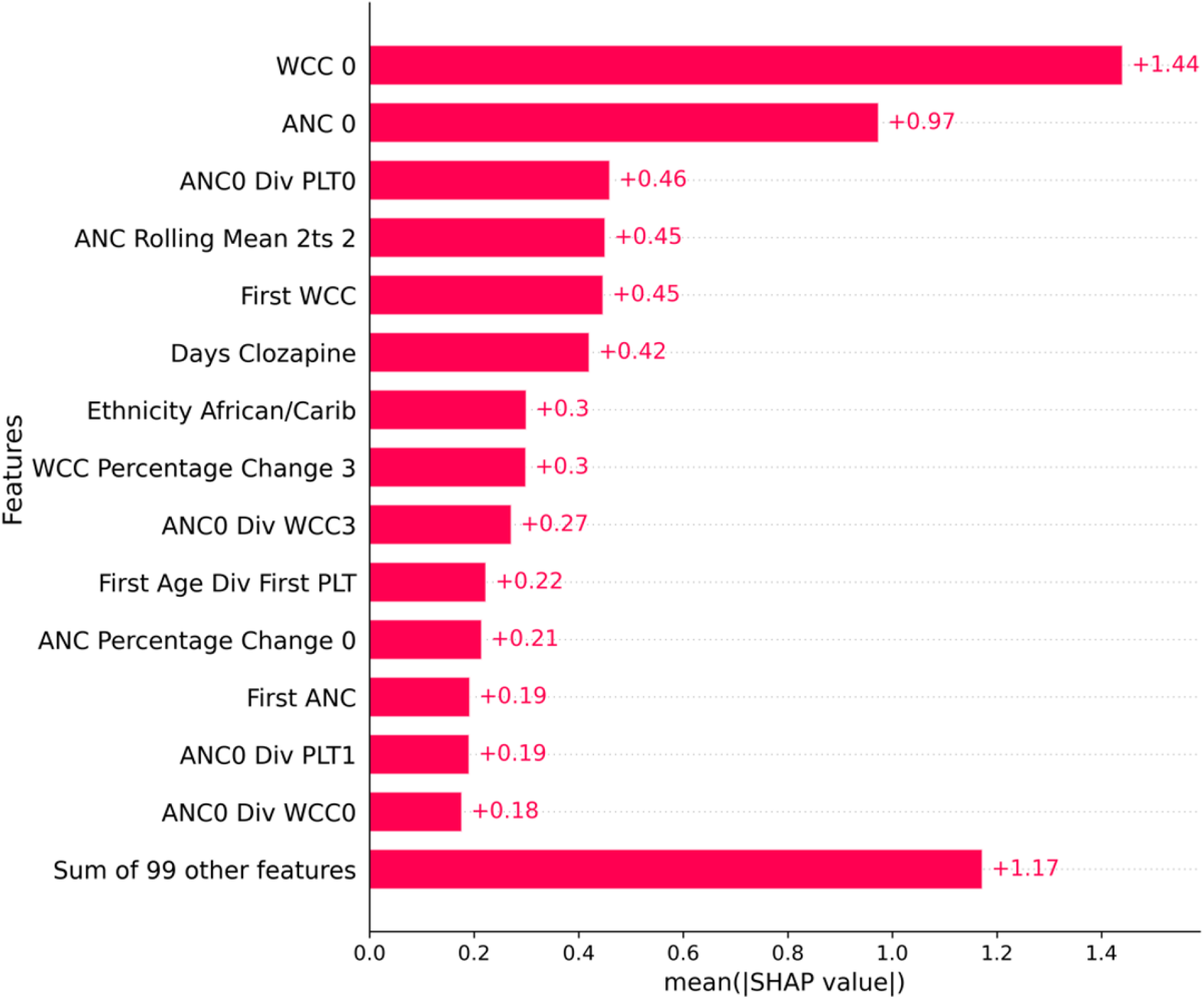
SHAP Analysis 1 week forecasting horizon

**Figure 13.**
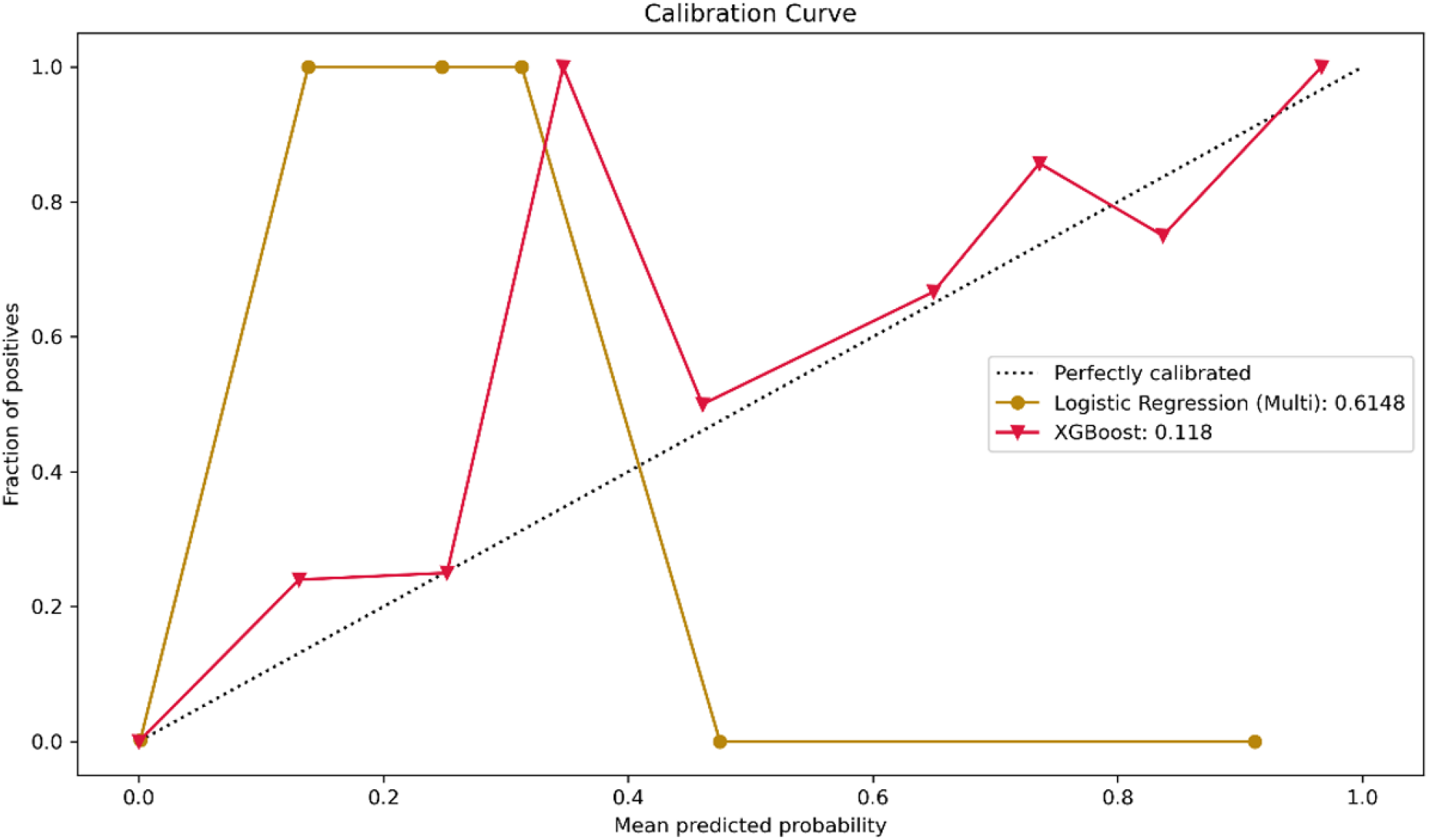
1-week forecasting horizon calibration curves for XGBoost and multivariate logistic regression models, showcasing calibration errors of 0.118 and 0.6148, respectively, without implementation of sample weights strategy.

**Figure 14.**
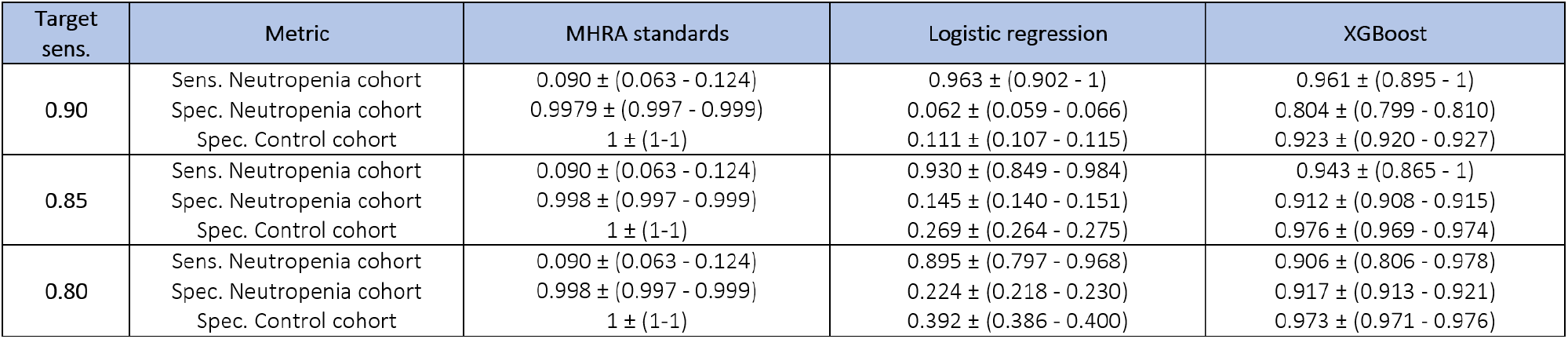
Predictive performance: 2 weeks forecasting horizon sensitivities and specificities

- Lower WCC and ANC at prediction time
- Lower ANC divided by PLT both at prediction time
- Higher ANC rolling mean from previous 3 weeks to previous 2 weeks
- Higher baseline WCC
- Short duration of clozapine treatment
- Negative for African/Caribbean ethnicity
- Lower mean WCC percentage change
- Lower ANC at prediction time divided by WCC from previous 4 weeks
- Older age divided by baseline PLT
- Higher baseline ANC
- Lower ANC at prediction time divided by PLT in previous week
- Lower ANC at prediction time divided by PLT in previous week
- Higher ANC at prediction time divided by WCC at prediction time

#### 2 weeks forecasting horizon

Figure 15 contains the SHAP values for the fortnightly model. Overall, and by order of importance, we can observe some of the features that are more likely to be associated with an event of CIA:

**Figure 15.**
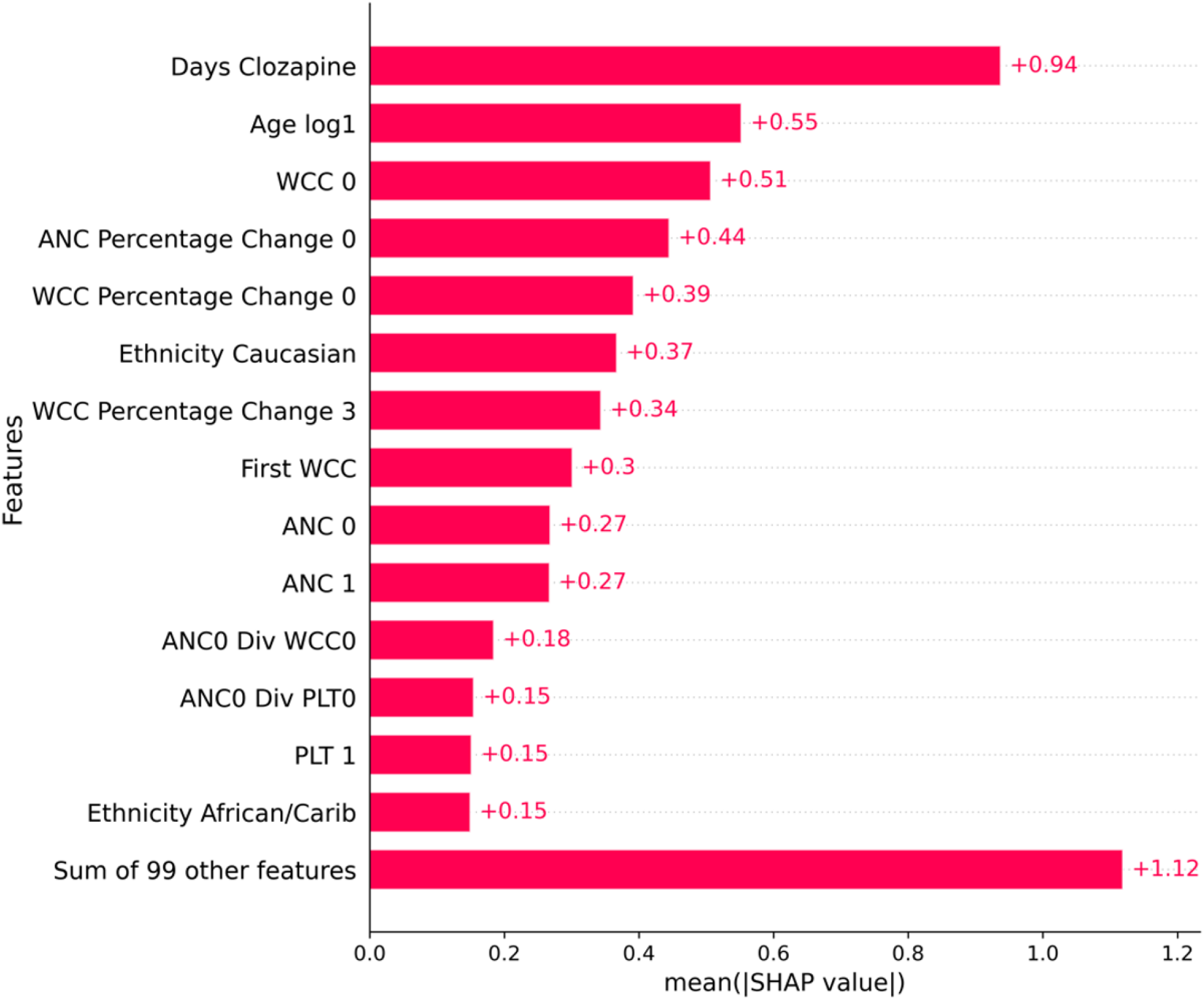
SHAP Analysis 2 weeks forecasting horizon

**Figure 16.**
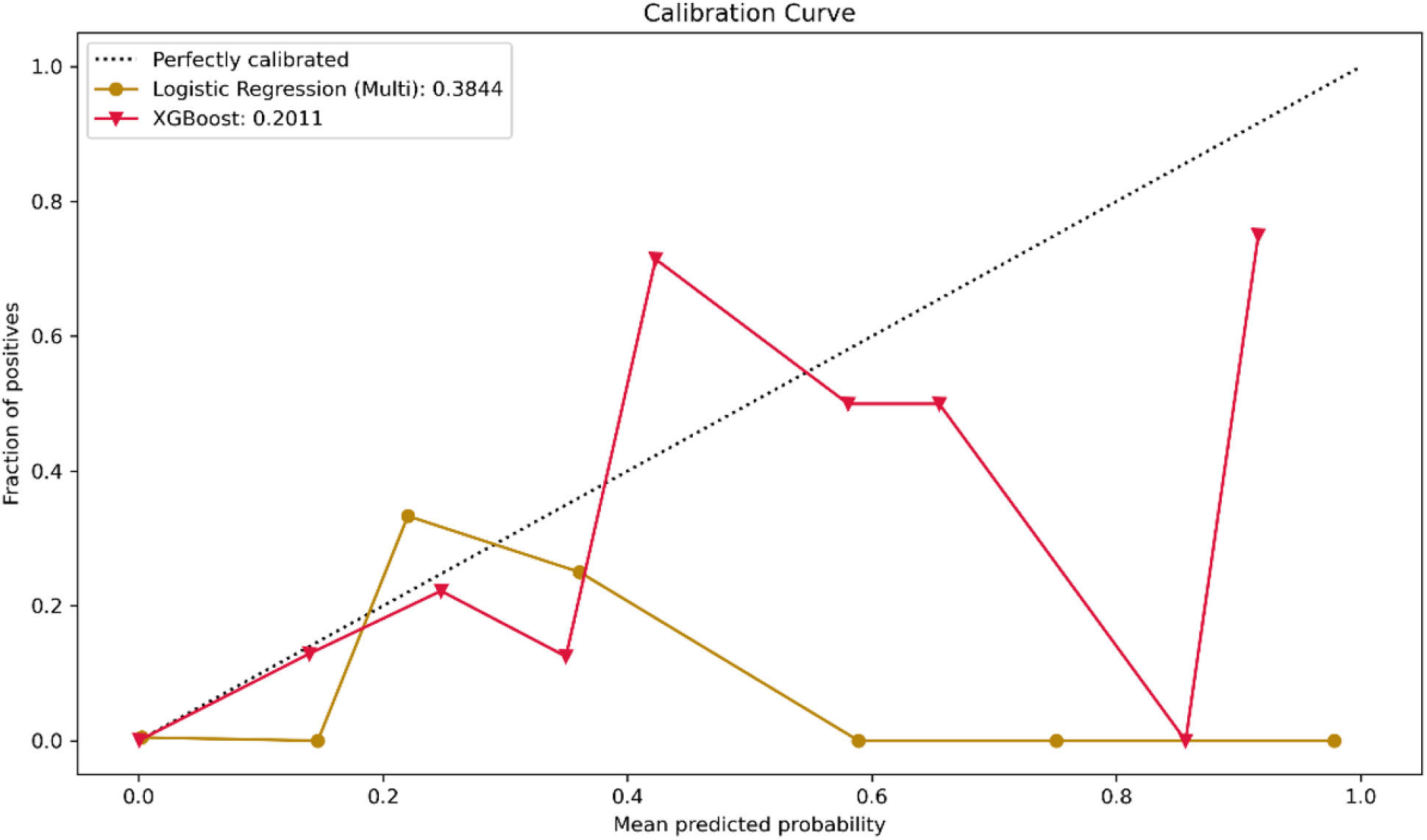
2 week forecasting horizon calibration curves for XGBoost and multivariate logistic regression models, showcasing calibration errors of 0.2011 and 0.3844, respectively, without implementation of sample weights strategy

**Figure 17.**
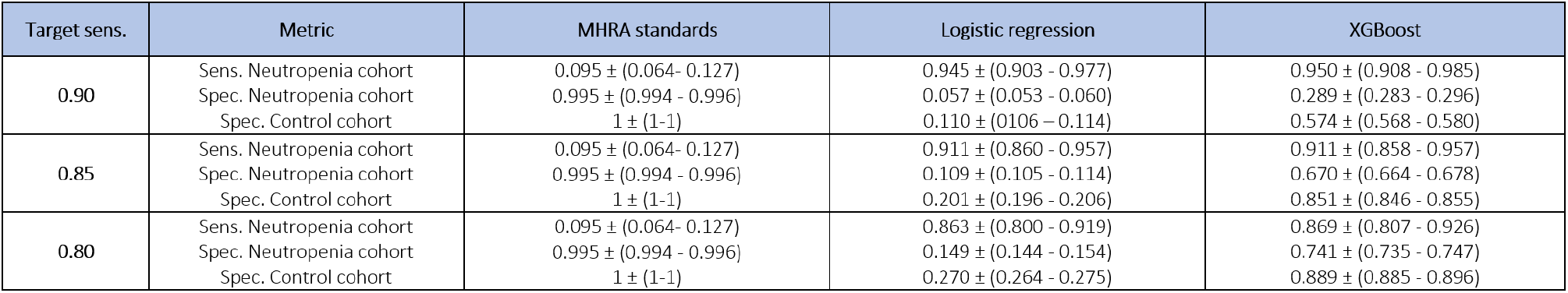
Predictive performance: 1 month forecasting horizon sensitivities and specificities

- Shorter duration of clozapine treatment
- Older age (logarithm)
- Lower WCC at prediction time
- Lower ANC percentage change at prediction time
- Lower WCC at prediction time
- Positive for Caucasian ethnicity
- Higher baseline WCC
- Lower ANC at prediction time and in previous 2 weeks
- Lower ANC at prediction time divided by WCC and PLT at prediction time
- Higher PLT in previous 2 weeks
- Negative for African/Caribbean ethnicity

#### 1 month forecasting horizon

Figure 18 contains the SHAP values for the monthly model. Overall, and by order of importance, we can observe some of the features that are more likely to be associated with an event of CIA:

**Figure 18.**
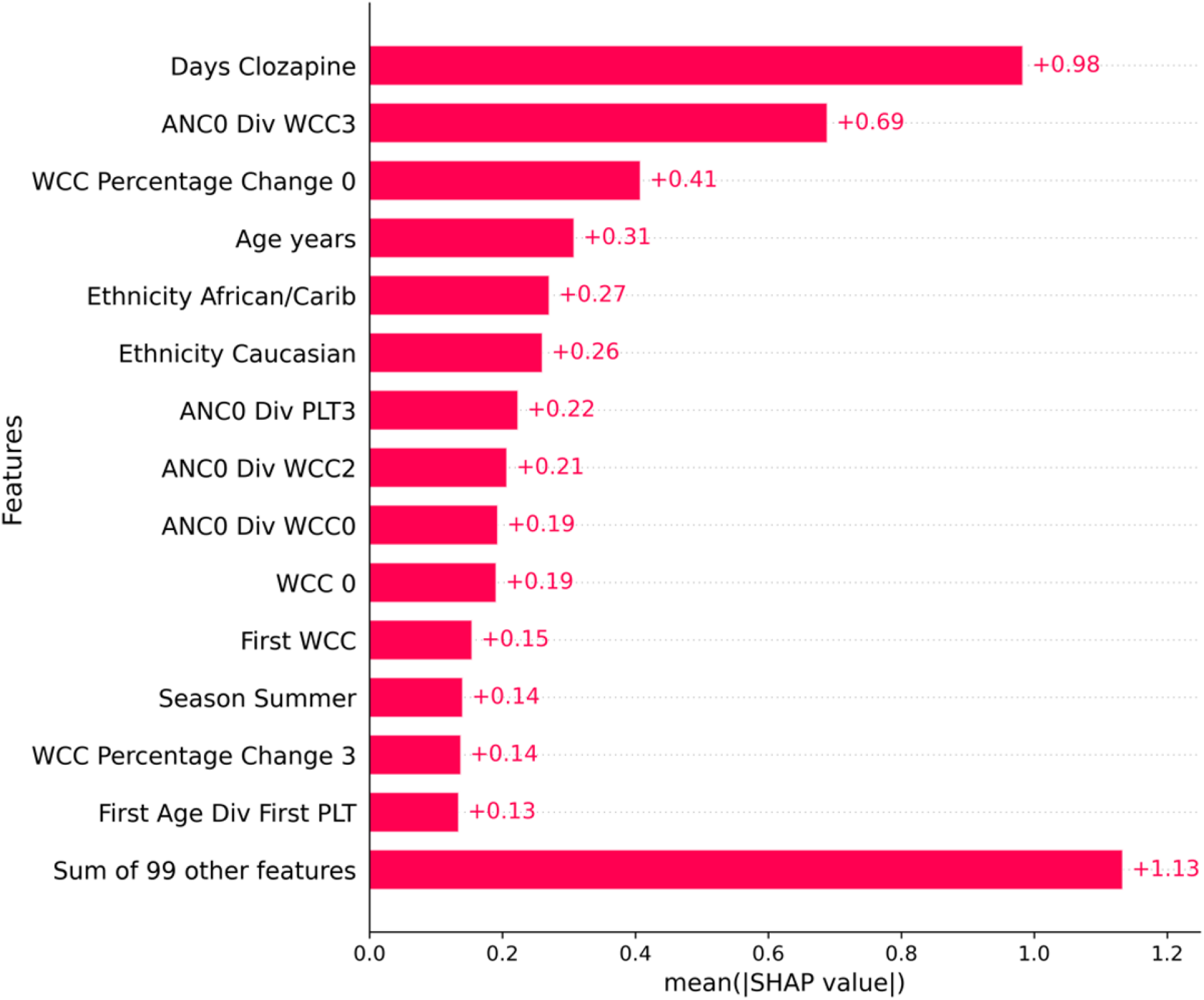
SHAP Analysis 1 month forecasting horizon

**Figure 19.**
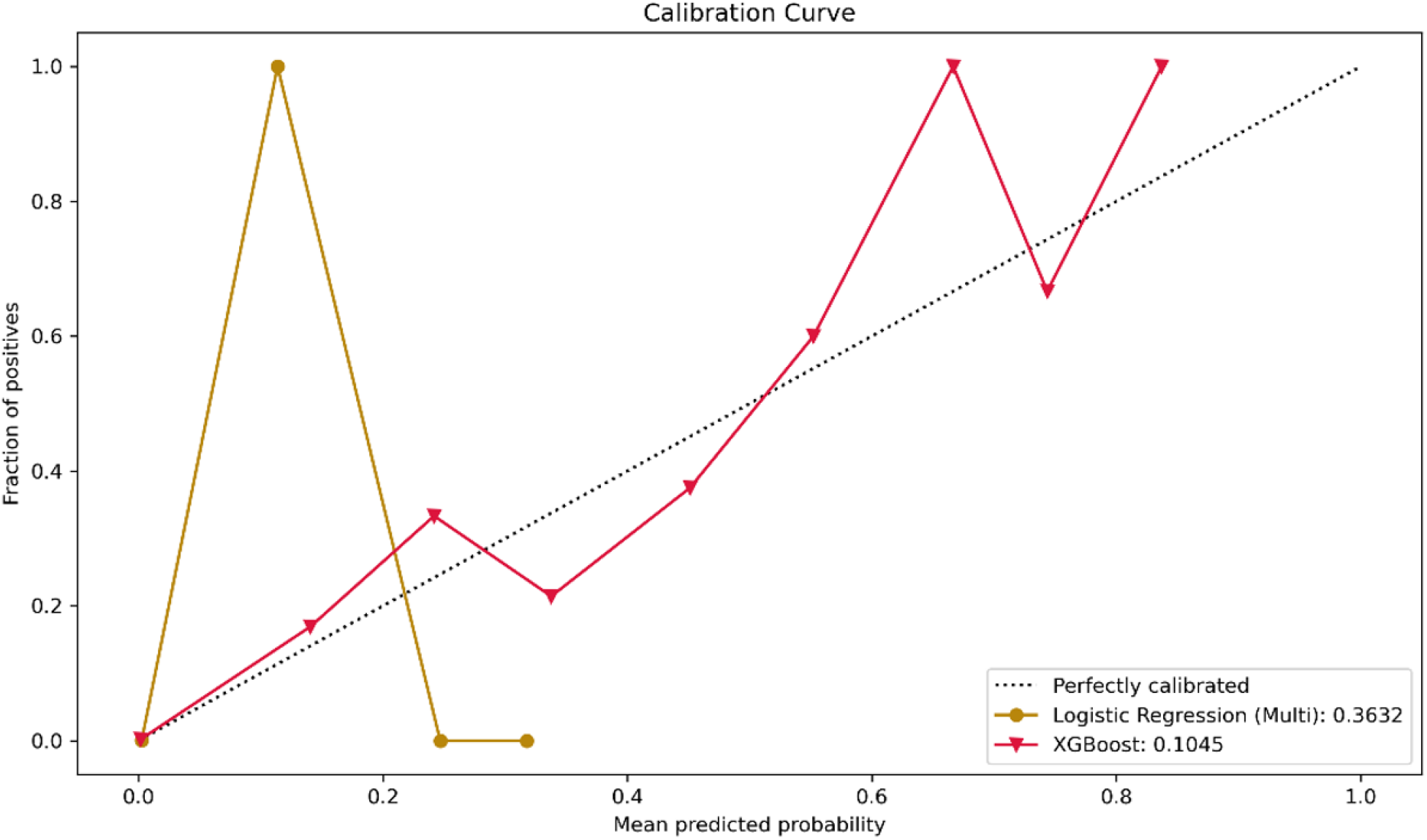
1 month forecasting horizon calibration curves for XGBoost and multivariate logistic regression models, showcasing calibration errors of 0.1045 and 0.3632, respectively, without implementation of sample weights strategy

**Figure 20.**
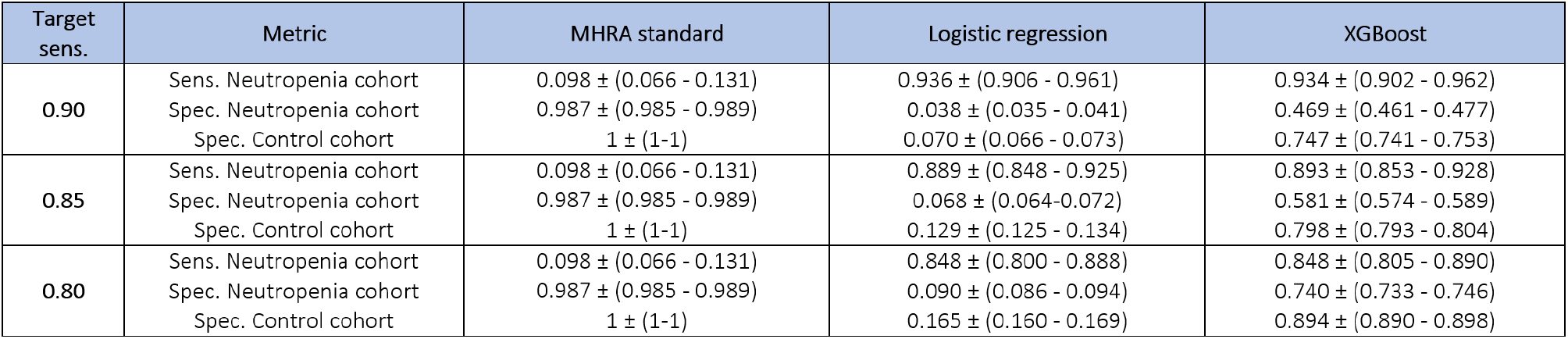
Predictive performance: 3 months forecasting horizon sensitivities and specificities

- Shorter duration of clozapine treatment
- Lower ANC at prediction time divided by WCC 3 months ago
- Lower WCC percentage change at prediction time from previous month
- Older age
- Negative for African/Caribbean ethnicity
- Positive for Caucasian ethnicity
- Low ANC value at prediction time divided by PLT values of 3 months ago
- Low ANC value at prediction time divided by WCC 2 months ago
- Low ANC value at prediction time divided by WCC at prediction time
- Higher baseline WCC
- Summer season
- Lower mean WCC percentage change
- Higher age divided by baseline PLT

#### 3 months forecasting horizon

Figure 21 contains the SHAP values for the 3 monthly model. Overall, and by order of importance, we can observe some of the features that are more likely to be associated with an event of CIA:

**Figure 21.**
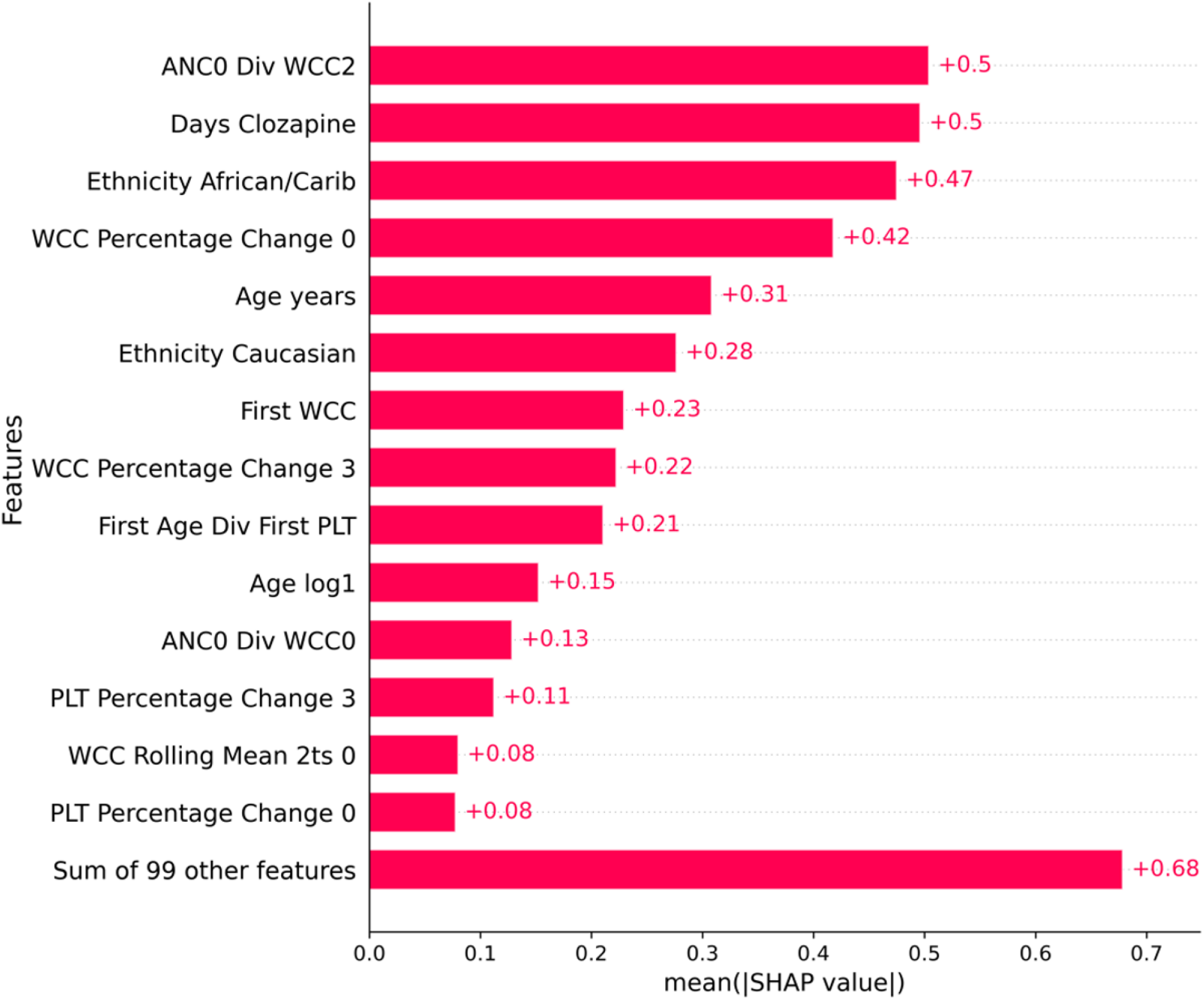
SHAP Analysis 3 months forecasting horizon

**Figure 22.**
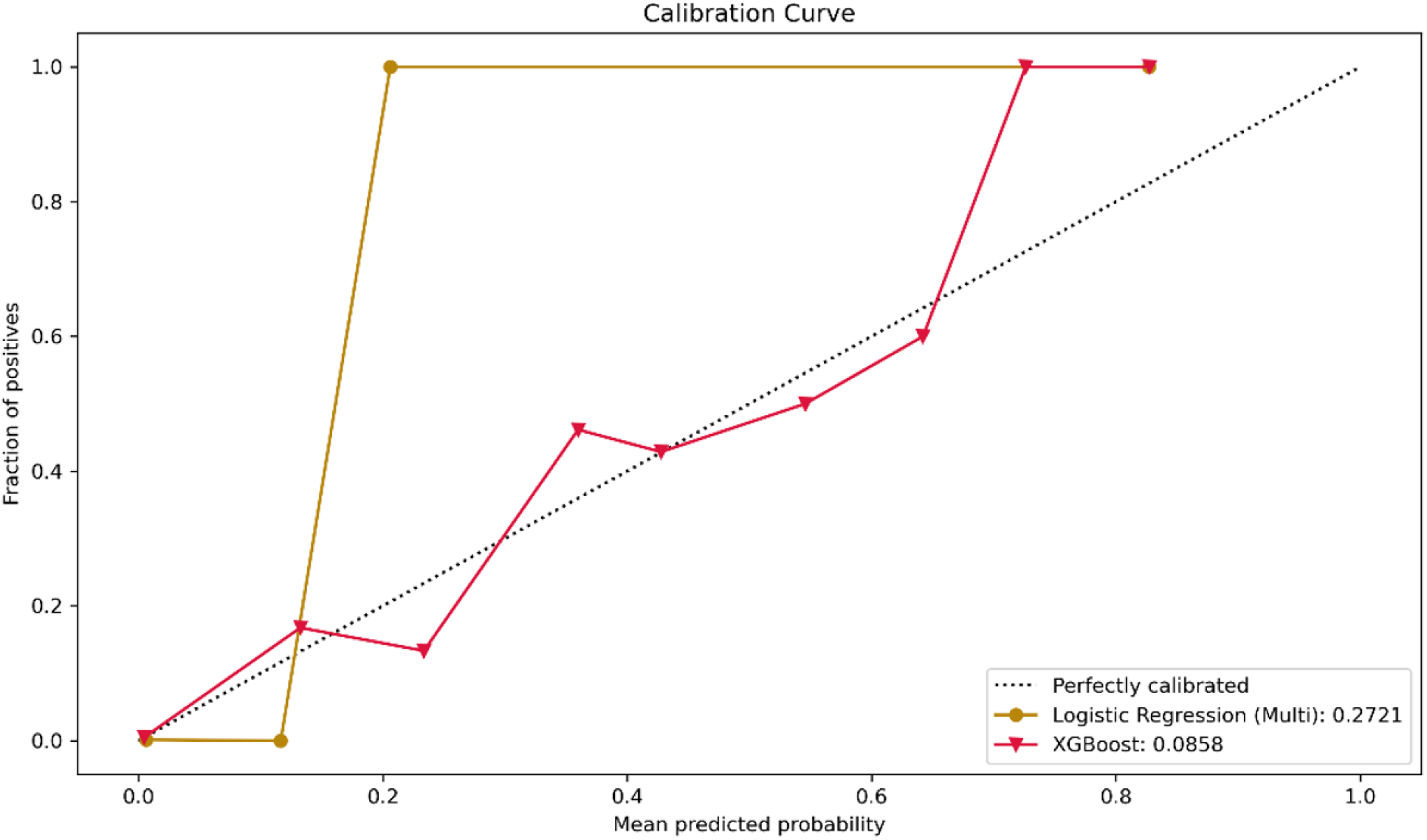
3 month forecasting horizon calibration curves for XGBoost and multivariate logistic regression models, showcasing calibration errors of 0.0858 and 0.2721, respectively, without implementation of sample weights strategy

**Figure 23.**
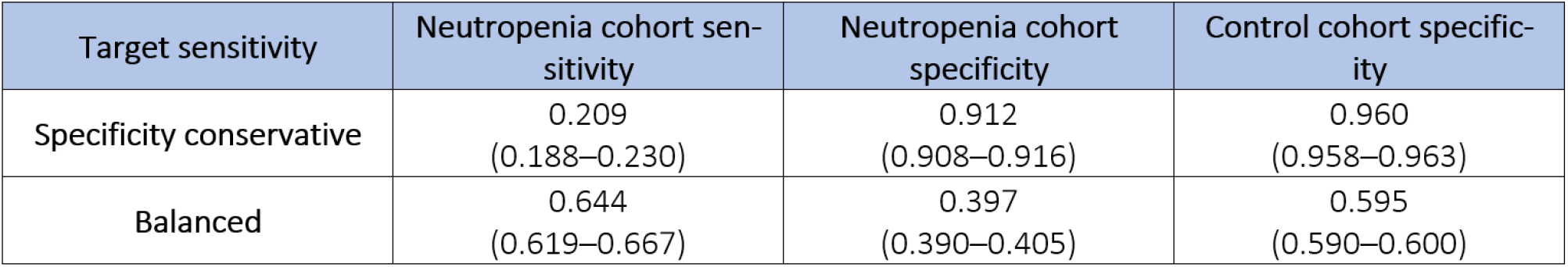
Baseline prediction model sensitivities and specificities. Target sensitivity thresholds for the specificity conservative and balanced (aiming to achieve high sensitivity) settings are 0.20 and 0.60, respectively

**Figure 24.**
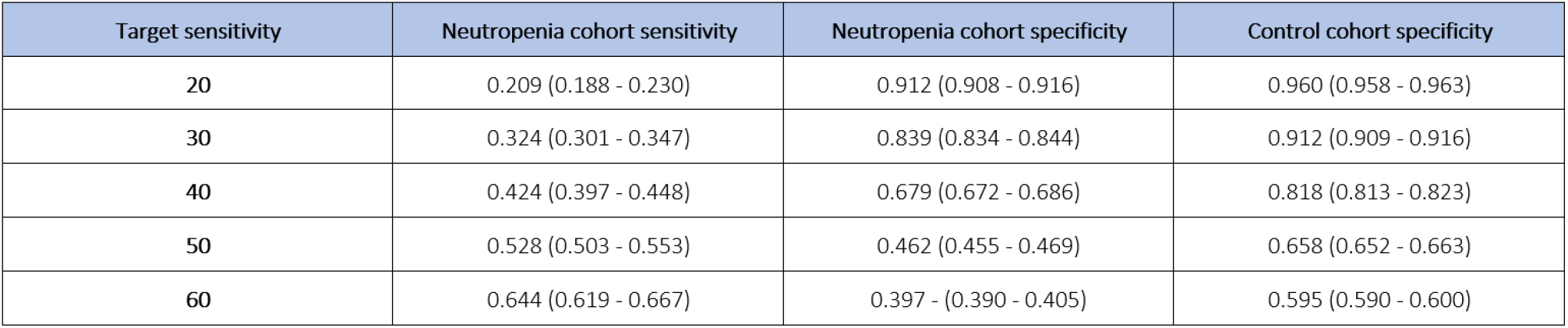
Baseline prediction model sensitivities and specificities at different target sensitivities

**Figure 25.**
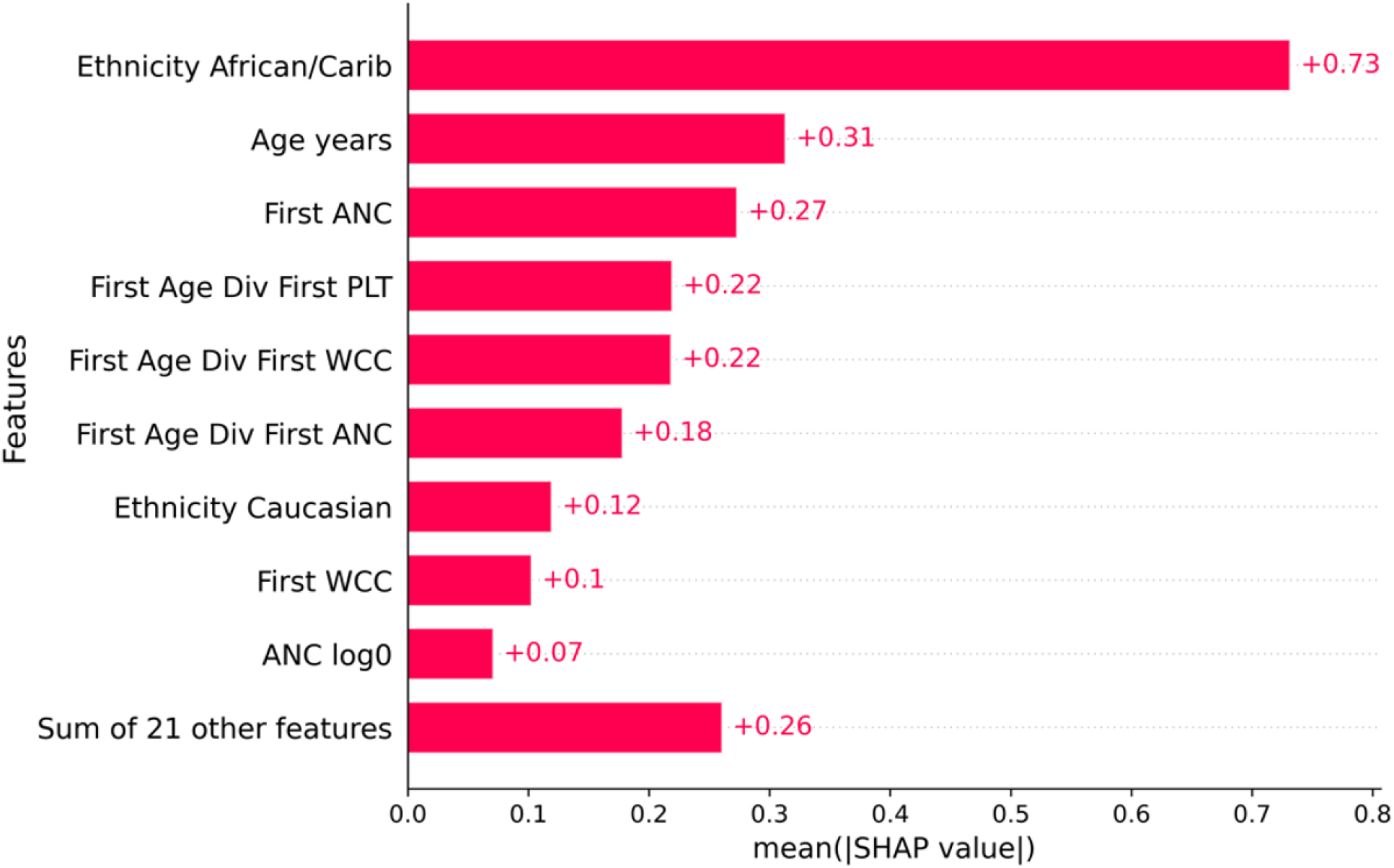
SHAP Analysis ML baseline prediction tool forecasting horizon

**Figure 26.**
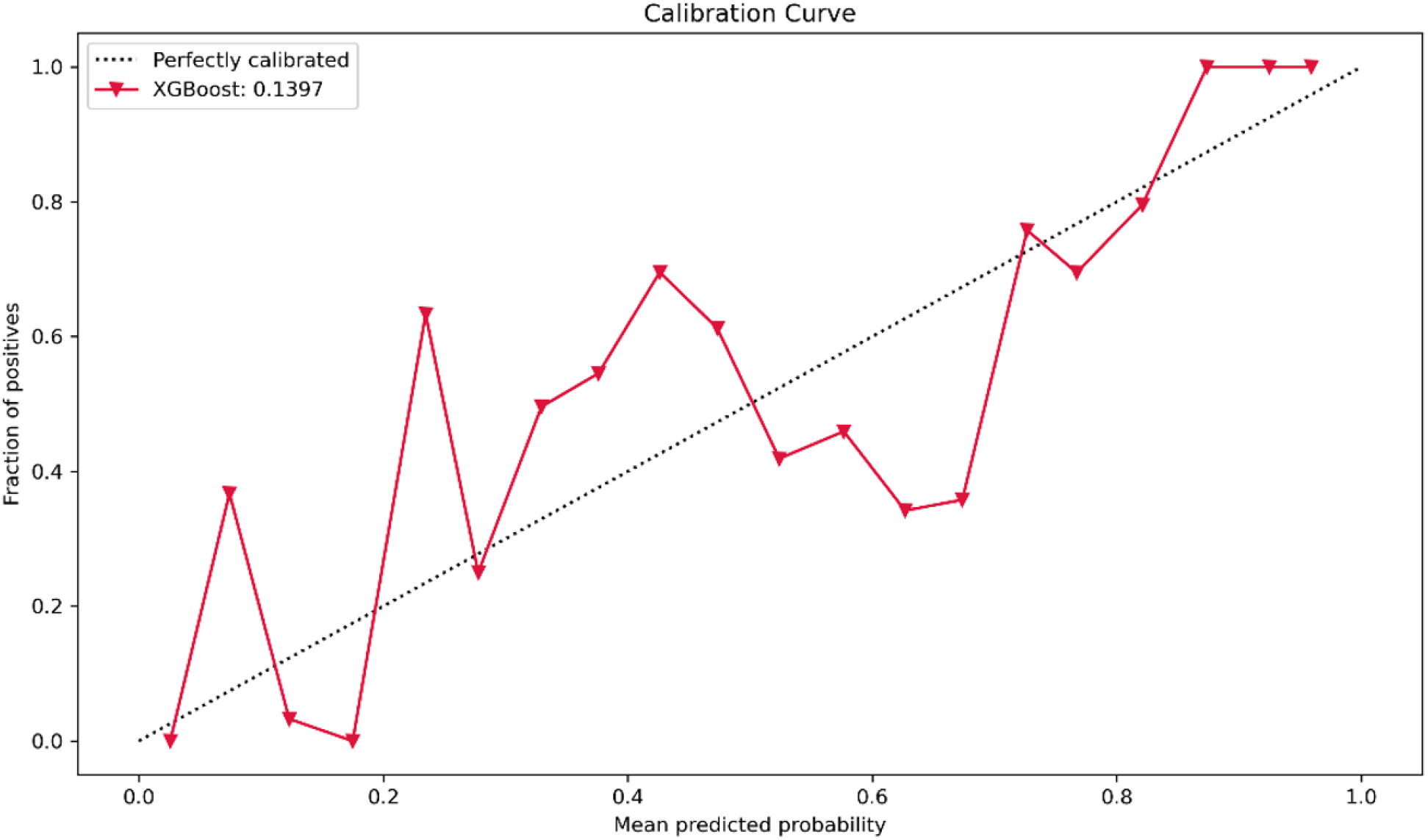
Profile-risk calibration curves for XGBoost model showcasing a calibration error of 0.1397.

**Figure 27.**
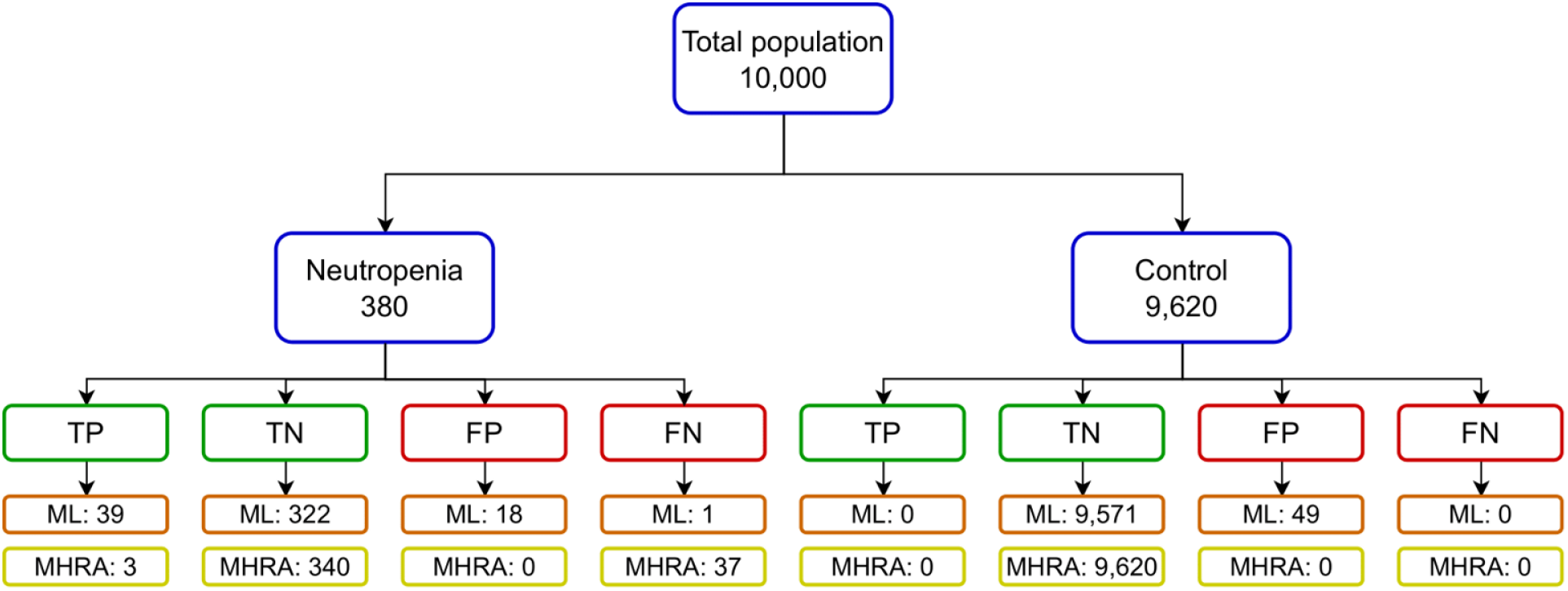
Practical exemplary comparison of ML and MHRA predictive performances across the 1-week forecasting scenario assuming previously investigated neutropenia and CIA expected ratios in a total population.

**Figure 28.**
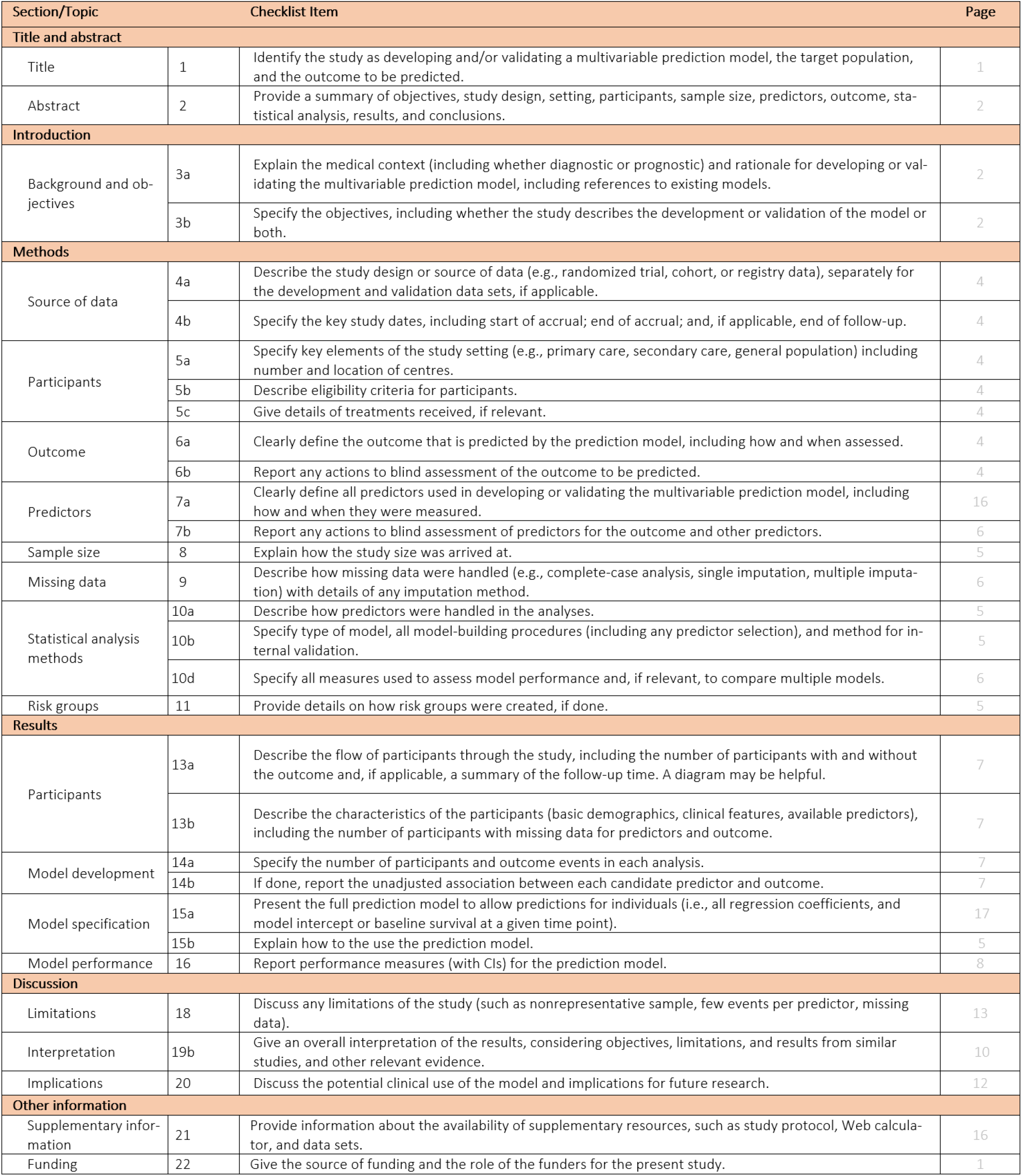
Transparent Reporting of a multivariable prediction model for Individual Prognosis or Diagnosis (TRIPOD) guidelines

- Low ANC at prediction time divided by WCC 6 months ago
- Shorter duration of clozapine treatment
- Positive for Caucasian ethnicity
- Negative for African/Caribbean ethnicity
- Low WCC percentage of change at prediction time from past
- Older age (raw or logarithm)
- Higher baseline WCC
- Lower mean WCC percentage change across the 4 measurements
- Higher baseline age divided by first PLT count
- Higher ANC divided by WCC at prediction time
- Higher percentage of PLT percentage change
- Lower WCC rolling mean from 9 months to 6 months ago
- Higher PLT percentage change at prediction time from the last period

#### Baseline risk assessment

Figure 25 contains the SHAP values for the weekly model. Overall, and by order of importance, we can observe some of the features that are more likely to be associated with an event of CIA:

- Negative for African/Caribbean ethnicity
- Older age
- Higher baseline ANC
- Older age divided by baseline PLT and WCC
- Lower age divided by baseline ANC
- Positive for Caucasian ethnicity
- Higher baseline WCC
- Higher logarithm ANC
- Negative for mixed ethnicity
- Lower ANC divided by PLT
- Higher baseline PLT
- Higher logarithm WCC
- Negative for TRS
- Male sex

#### TRIPOD

